# THE US MIDLIFE MORTALITY CRISIS CONTINUES: EXCESS CAUSE-SPECIFIC MORTALITY DURING 2020

**DOI:** 10.1101/2021.05.17.21257241

**Authors:** Dana A. Glei

**Author notes:** **Address correspondence to:** Dana A. Glei, 5985 San Aleso Court, Santa Rosa, CA 95409.

## Abstract

COVID-19 prematurely ended many lives, particularly among the oldest Americans, but the pandemic also had an indirect effect on health and non-COVID mortality among the working-age population, who suffered the brunt of the economic consequences. This analysis investigates whether monthly excess mortality in the US during 2020 varied by age and cause of death. Based on national-level death counts and population estimates for 1999-2020, negative binomial regression models—fit separately by sex— were used to estimate monthly cause-specific excess mortality by age group during 2020. Among males, 71% non-COVID excess deaths occurred at working ages (25-64), but those ages accounted for only 36% of non-COVID excess deaths in females. The results revealed substantial numbers of excess deaths from external causes (particularly among males), heart disease, diabetes, Alzheimer’s disease (particularly among women), and cerebrovascular disease. For males, the largest share of non-COVID excess deaths resulted from external causes, nearly 80% of which occurred at working ages. Although incorrectly classified COVID-19 deaths may explain some excess non-COVID mortality, misclassification is unlikely to explain the increase in external causes of mortality. Auxiliary analyses suggested that drug-related mortality may be driving the rise in external mortality, but drug overdoses were already increasing for a full year prior to the pandemic. The oldest Americans bore the brunt of COVID-19 mortality, but working-age Americans, particularly men, suffered substantial numbers of excess non-COVID deaths, most commonly from external causes and heart disease.

## INTRODUCTION

Older Americans suffered the highest COVID-19 mortality, but we may be underestimating the indirect impact of the pandemic on mortality from other causes among the working-age population. Only 38% of the excess mortality^*^ among Americans aged 25-44 during March-July 2020 could be attributed directly to COVID-19.^1^

Although much less likely than older Americans to die from COVID-19, younger Americans bore the brunt of the economic consequences as their work lives were abruptly altered. Essential workers continued to be in high demand, but faced additional stressors (e.g., fear of exposure to the virus; shortages of personal protective equipment). An additional threat was posed by recalcitrant individuals who resisted public health orders, particularly when enforced by low-wage workers (e.g., grocery/retail store employees). Business owners had to grapple with logistical challenges resulting from new public health regulations and the financial aftermath of reduced demand for their services. Other workers suffered sudden income decline because of job loss or reduced hours. The most fortunate workers retained their jobs with reduced exposure to SARS-CoV-2, but had to adapt to working from home and learning new technologies.

The unexpected closure of child care centers and schools further exacerbated the conflicts between work and family life for young and midlife Americans. With no warning, parents of young children lost access to child care services and became responsible for helping to homeschool their children, while continuing to juggle work demands and the potential needs of their own parents, who were particularly vulnerable to COVID-19.

Depending on the period of coverage and methodology, prior studies have reported 16-23% excess mortality during the pandemic,^2–4^ but only 72-89% of the excess deaths could be attributed directly to COVID-19.^2–5^ During the period from January 26, 2020 to October 3, 2020, Americans aged 25-44 suffered the largest relative increase in excess mortality (27% peaking at nearly 50% in mid-July), whereas those aged 85 and older experienced a smaller increment (15%, but also peaking near 50% in late April).^6^ Excess mortality peaked earlier—in April—and declined more rapidly for Americans aged 45 and older than for their younger counterparts (aged 25-44), among whom excess mortality was persistently high (>30%) throughout early April to early August, peaking in mid-July.^6^

It remains unclear how excess mortality from causes <u>other than COVID-19</u> is distributed by cause and the extent to which it varies by sex and age. Compared with 2019, the absolute number of deaths in 2020 increased for various causes: diabetes (15%), unintentional injuries (11%), Alzheimer’s (10%), stroke (6%), influenza & pneumonia (6%), and heart disease (5%).^7^ That same report showed virtually no change in the number of deaths from cancer or kidney disease and revealed a small decline in deaths from chronic lower respiratory diseases (−3%) and suicide (−6%). Other evidence suggested that mortality from drug overdoses,^8^ homicides,^9^ and motor vehicle accidents^10^ also increased during the pandemic. The extent of change in alcohol-related mortality for the US as a whole remains unknown, but some evidence showed increased alcohol consumption, particularly among women and Americans aged 30-59,^11^ and provisional data for Minnesota implied that alcohol-attributable deaths were 25-65% higher in June-December 2020 than in the same months in 2019.^12^ To my knowledge, no US study has investigated the extent to which excess mortality for specific causes of death varies by sex and age.

This analysis quantifies the changes in mortality for selected causes of death during the COVID-19 pandemic up to December 31, 2020. In addition, the models evaluate the extent to which the levels of excess mortality varied by sex, age group, and across the months of 2020.

## METHODS

### Data

These analyses were based on national-level monthly death counts by sex, age group (0-4, 5-14, 15-24, 25-34,…75-84, 85+), and selected causes of death for the period from January 1999 to December 2020 (see eText 1 for more details).^13,14^ Although the provisional data covered the period through June 2021,^14^ this analysis was restricted to the period through December 31, 2020, because the data for 2021 are likely to be incomplete.^15^ Annual mid-year population estimates by sex and age^16,17^ were used to compute the cause-specific death rates. Collectively, the dataset represented 56.8 million deaths over 6.7 billion person-years of observation for the entire US population.

### Measures

#### Outcome: Age-Specific Death Rates for Selected Causes of Death

The first model comprised all-cause mortality (including deaths from COVID-19). Next, models were fit separately for 8 groups of underlying causes (see eText 2 for details).

#### Control Variables

To account for the pre-pandemic patterns of mortality during 1999-2019, the models controlled for age, calendar year, and seasonality. The specification of the time trend varied by outcome (see Analytical Strategy).

#### Mortality During 2020

The key variables of interest were the dichotomous indicators for each month during 2020. To test whether excess mortality differed by age, a set of dummy variables for broad age groups (i.e., <15, 15-24, 25-44, 45-64, 65-74, 75+) was interacted with a dummy variable for the pandemic period (March-December 2020).

### Analytical Strategy

The monthly age-specific mortality rates for each cause of death were modeled separately by sex using a negative binomial model (see eText 3 for the detailed equation). The model included control variables to account for the age pattern, seasonality, and the pre-pandemic time trend. Because the seasonality of mortality can vary by age,^18^ interactions between month and age were included in the models for all-cause mortality and external causes. Those interactions did not improve model fit for the remaining outcomes and thus, were omitted. The functional form for the time trend varied by outcome to maximize predictive accuracy in 2019: linear specification for influenza/pneumonia mortality; quadratic specification for all-cause, heart disease, cerebrovascular disease, Alzheimer’s disease, diabetes, and external causes; cubic specification for other respiratory diseases (excluding COVID-19) and cancer. All models included interactions between the time trend and age because recent mortality decline has varied by age.^19^

An additional set of dummy variables for each month of 2020 were included to estimate the extent to which monthly mortality differed from the pre-pandemic time trend. Prior to March, there were few COVID-19 deaths in the US, and the pandemic had yet to make a substantial impact on everyday life here. Thus, the excess mortality rate ratios were expected to be close to 1.0 for January and February. Interactions between broad age groups and a dichotomous indicator for the pandemic (March-December) period of 2020 were used to test for age differences in excess mortality.

To evaluate the sensitivity of the results to the specification of the pre-pandemic time trend, an auxiliary set of models were estimated using alternative specifications for the time trend. Additional models assessed the sensitivity to the length of the time series used to fit the model by restricting the data series to: a) the period since 2009; and b) the period since 2014.

Finally, auxiliary models were estimated to test whether excess mortality varied by both age group and wave of the pandemic. Those additional interactions were omitted from the final analysis because they did not improve model fit based on the Bayesian Information Criterion (BIC).

For some groups of causes, the death rates were exceptionally low at young ages, which can result in very high rate ratios. Thus, the cause-specific models were restricted to the age range within which the average death rates in each age group across 1999-2020 were at least one per million: ages 15 and older for heart diseases; ages 25 and older for other respiratory diseases, cerebrovascular diseases, and diabetes mellitus; ages 35 and older for influenza/pneumonia; and ages 55 and older for Alzheimer’s disease.

The estimated number of excess deaths was computed as the difference between: 1) the predicted number of deaths in 2020 based on the model coefficients, and 2) the predicted number of deaths when the 2020 monthly parameters and the interactions between age and the pandemic indicator was reset to zero (i.e., “expected” deaths in the absence of a pandemic). The estimated number of excess non-COVID deaths was derived by subtracting the reported number of COVID-19 deaths from the estimated number of excess deaths from all causes.

## RESULTS

Death rates during January and February 2020 were generally similar to or lower than expected based on the pre-pandemic trends with one major exception: mortality from influenza/pneumonia was notably higher (eTables 1-3). Throughout April-December 2020, all-cause death rates above age 15 were substantially higher than expected (Figure 1). In relative terms, excess mortality from all causes was similar across ages 15 and older, but the absolute number of excess deaths was much larger at the oldest ages (Figure 2). At younger ages, where mortality rates are low, a substantial increase in the relative risk can translate into a very small absolute increment in the number of deaths. For example, the excess mortality ratio ratios at ages 25-34 were as high, if not higher, than at ages 64-74, but the estimated number of excess deaths in March-December 2020 was much lower for those aged 25-34 (21,300 males; 8,260 females) than for those 64-74 (54,284 and 36,815, respectively; Tables 1 and 2). Americans aged 65 and older accounted for 64% of the 252,199 excess deaths in males (Table 1) and 81% of the 196,481 excess deaths in females (Table 2).

**Figure 1.**
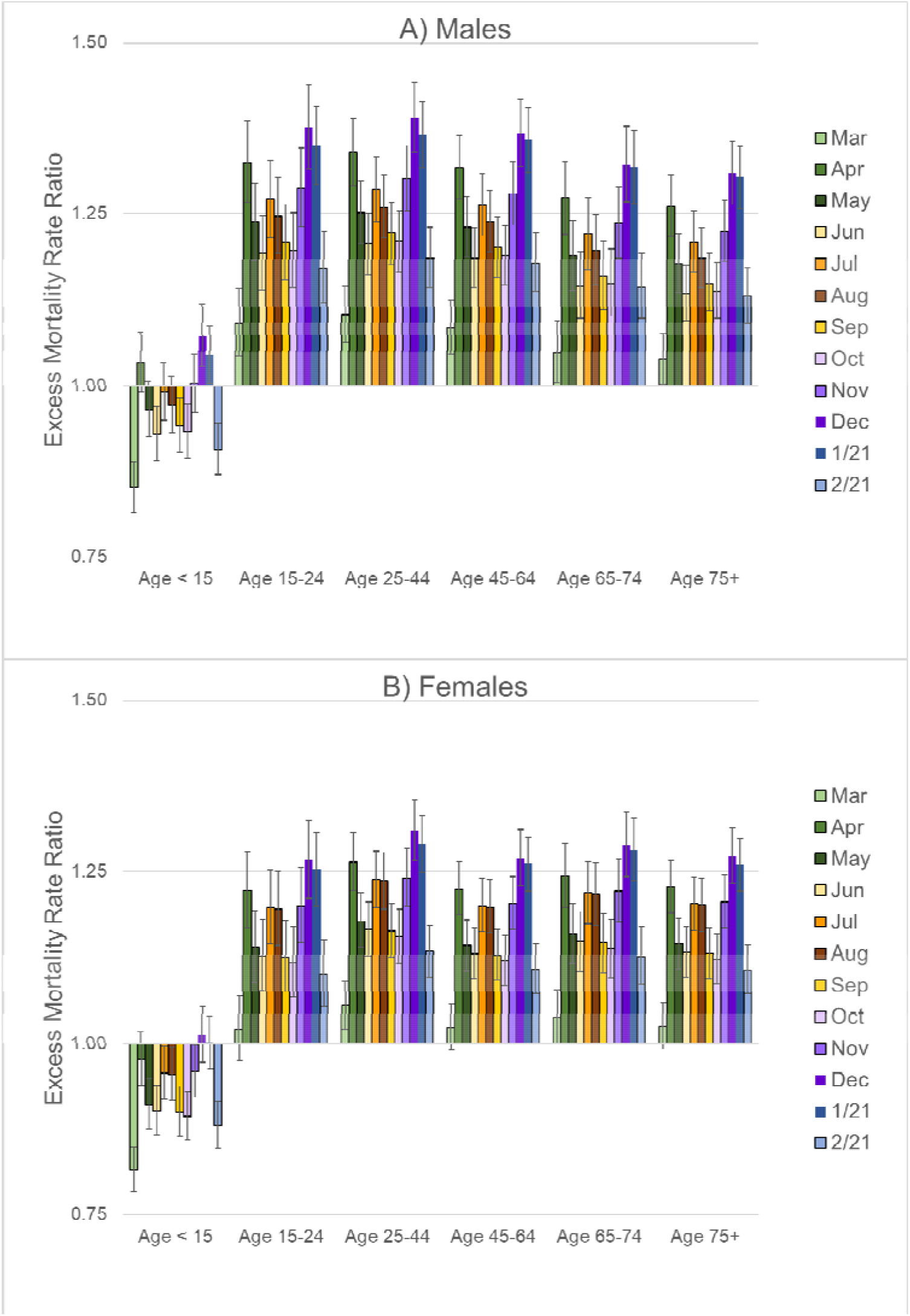
Excess Mortality Rate Ratios from All-Cause Mortality (Including COVID-19) by Age Group During March-December 2020, US: A) Males, B) Females. Note: These estimates are based on the regression models shown in eTable 1. A rate ratio higher than 1.0 implies that the mortality rates were higher than expected based on the pre-pandemic trends, whereas a value less than 1.0 suggest lower than expected mortality rates. The bars on the graph of excess mortality rate ratios represent the 95% confidence interval.

**Figure 2.**
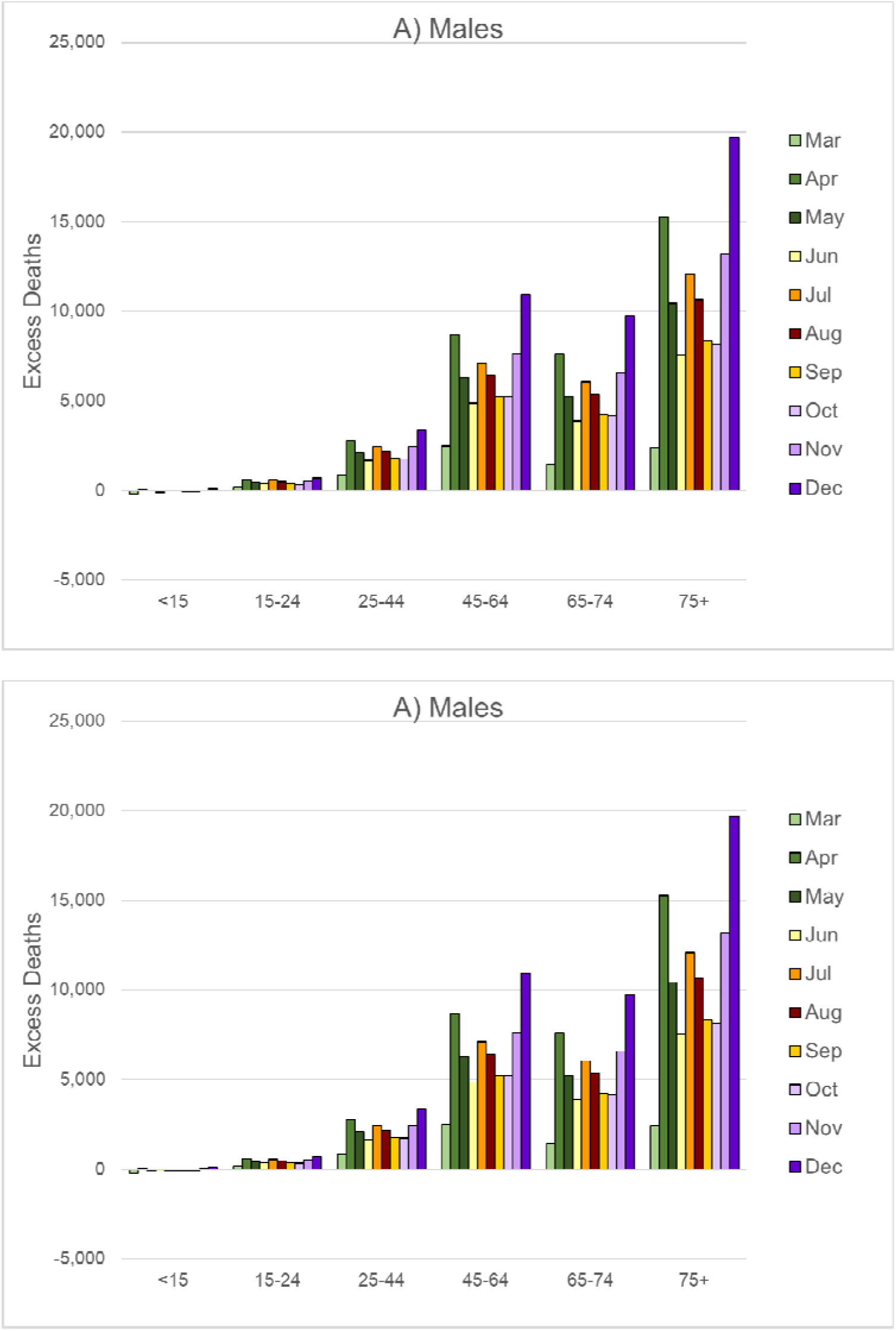
Number of Excess Deaths from All-Cause Mortality (Including COVID-19) by Age Group During March-December 2020, US: A) Males, B) Females. Note: These estimates are based on the regression models shown in eTable 1.

**Table 1.** Estimated Number and Percentage of Excess Deaths During March-December 2020 by Age Group and Cause, Males.

|  | Total | Age<15 | Age 15-24 | Age 25-44 | Ages 45-64 | Ages 65-74 | Ages 75+ |
| --- | --- | --- | --- | --- | --- | --- | --- |
| All causes, N | 252,199 | -446 | 4,495 | 21,300 | 64,862 | 54,284 | 107,703 |
| % of Total across all ages | 100.0% | -0.2% | 1.8% | 8.4% | 25.7% | 21.5% | 42.7% |
| COVID-19 (underlying cause), N | 192,928 | 62 | 309 | 5,552 | 38,555 | 47,137 | 101,313 |
| % of Total across all ages | 100.0% | 0.0% | 0.2% | 2.9% | 20.0% | 24.4% | 52.5% |
| % of Total excess deaths | 76.5% | -13.9% | 6.9% | 26.1% | 59.4% | 86.8% | 94.1% |
| All causes excluding COVID-19, N<sup>a</sup> | 59,271 | -508 | 4,186 | 15,748 | 26,307 | 7,147 | 6,390 |
| % of Total across all ages | 100.0% | -0.9% | 7.1% | 26.6% | 44.4% | 12.1% | 10.8% |
| <u>Modeled separately by underlying cause</u> |  |  |  |  |  |  |  |
| Influenza/pneumonia, N | 2,441 | b | b | 64<sup>c</sup> | 682 | 1,100 | 595 |
| % of Total across ages 35+ | 100.0% | b | b | 2.6% | 27.9% | 45.1% | 24.4% |
| Other respiratory disease, N | -1,150 | b | b | 228 | 591 | -300 | -1,669 |
| % of Total across ages 25+ | 100.0% | b | b | -19.8% | -51.4% | 26.1% | 145.2% |
| Heart disease, N | 15,943 | b | 0 | 1,589 | 7,122 | 2,534 | 4,698 |
| % of Total across ages 15+ | 100.0% | b | 0.0% | 10.0% | 44.7% | 15.9% | 29.5% |
| Cerebrovascular disease, N | 2,881 | b | b | 187 | 1,121 | 615 | 958 |
| % of Total across ages 25+ | 100.0% | b | b | 6.5% | 38.9% | 21.3% | 33.2% |
| Cancer, N | 266 | -24 | 9 | 41 | 672 | 73 | -506 |
| % of Total across all ages | 100.0% | -8.9% | 3.3% | 15.5% | 252.6% | 27.6% | -190.1% |
| Alzheimer's disease, N | 2,046 | b | b | b | 24<sup>d</sup> | 247 | 1,775 |
| % of Total across ages 55+ | 100.0% | b | b | b | 1.2% | 12.1% | 86.8% |
| Diabetes, N | 7,292 | b | b | 535 | 2,183 | 1,947 | 2,626 |
| % of Total across ages 25+ | 100.0% | b | b | 7.3% | 29.9% | 26.7% | 36.0% |
| External causes, N | 17,341 | 189 | 3,764 | 8,959 | 4,869 | 76 | -515 |
| % of Total across all ages | 100.0% | 1.1% | 21.7% | 51.7% | 28.1% | 0.4% | -3.0% |

<sup>a</sup> Derived by subtracting the reported number of COVID-19 deaths (as the underlying cause) from the estimated number of excess deaths from all causes.
<sup>b</sup> These ages were excluded from the model because mortality rates from this cause are extremely low.
<sup>c</sup> The model was restricted to ages 35 and older because mortality rates from this cause are exceptionally low below age 35.
<sup>d</sup> The model was restricted to ages 55 and older because there are very few deaths from Alzheimer's disease below age 55.

**Table 2.** Estimated Number and Percentage of Excess Deaths During March-December 2020 by Age Group and Cause, Females.

|  | Total | Age<15 | Age 15-24 | Age 25-44 | Ages 45-64 | Ages 65-74 | Ages 75+ |
| --- | --- | --- | --- | --- | --- | --- | --- |
| **Number of excess deaths** |  |  |  |  |  |  |  |
| All causes, N | 196,481 | -798 | 1,113 | 8,260 | 29,047 | 36,815 | 122,045 |
| % of Total across all ages | 100.0% | -0.4% | 0.6% | 4.2% | 14.8% | 18.7% | 62.1% |
| COVID-19 (underlying cause), N | 158,485 | 41 | 196 | 2,848 | 20,701 | 29,296 | 105,403 |
| % of Total across all ages | 100.0% | 0.0% | 0.1% | 1.8% | 13.1% | 18.5% | 66.5% |
| % of Total excess deaths | 80.7% | -5.1% | 17.6% | 34.5% | 71.3% | 79.6% | 86.4% |
| All causes excluding COVID-19, N<sup>a</sup> | 37,996 | -839 | 917 | 5,412 | 8,346 | 7,519 | 16,642 |
| % of Total across all ages | 100.0% | -2.2% | 2.4% | 14.2% | 22.0% | 19.8% | 43.8% |
| <u>Modeled separately by underlying cause</u> |  |  |  |  |  |  |  |
| Influenza/pneumonia, N | -54 | <sup>b</sup> | <sup>b</sup> | 22<sup>c</sup> | 166 | 487 | -729 |
| % of Total across ages 35+ | 100.0% | <sup>b</sup> | <sup>b</sup> | -41.9% | -310.3% | -909.9% | 1362.1% |
| Other respiratory disease, N | -5,433 | <sup>b</sup> | <sup>b</sup> | 87 | 115 | -1,037 | -4,598 |
| % of Total across ages 25+ | 100.0% | <sup>b</sup> | <sup>b</sup> | -1.6% | -2.1% | 19.1% | 84.6% |
| Heart disease, N | 10,566 | <sup>b</sup> | 21 | 607 | 2,110 | 2,294 | 5,535 |
| % of Total across ages 15+ | 100.0% | <sup>b</sup> | 0.2% | 5.7% | 20.0% | 21.7% | 52.4% |
| Cerebrovascular disease, N | 2,440 | <sup>b</sup> | <sup>b</sup> | 74 | 538 | 694 | 1,134 |
| % of Total across ages 25+ | 100.0% | <sup>b</sup> | <sup>b</sup> | 3.0% | 22.1% | 28.4% | 46.5% |
| Cancer, N | -2,429 | -17 | -5 | -330 | -821 | 675 | -1,931 |
| % of Total across all ages | 100.0% | 0.7% | 0.2% | 13.6% | 33.8% | -27.8% | 79.5% |
| Alzheimer's disease, N | 7,716 | <sup>b</sup> | <sup>b</sup> | <sup>b</sup> | 14<sup>d</sup> | 685 | 7,018 |
| % of Total across ages 55+ | 100.0% | <sup>b</sup> | <sup>b</sup> | <sup>b</sup> | 0.2% | 8.9% | 90.9% |
| Diabetes, N | 6,433 | <sup>b</sup> | <sup>b</sup> | 279 | 1,149 | 1,551 | 3,455 |
| % of Total across ages 25+ | 100.0% | <sup>b</sup> | <sup>b</sup> | 4.3% | 17.9% | 24.1% | 53.7% |
| External causes, N | 3,536 | 10 | 638 | 2,605 | 1,150 | -278 | -589 |
| % of Total across all ages | 100.0% | 0.3% | 18.0% | 73.7% | 32.5% | -7.9% | -16.6% |

<sup>a</sup> Derived by subtracting the reported number of COVID-19 deaths (as the underlying cause) from the estimated number of excess deaths from all causes.
<sup>b</sup> These ages were excluded from the model because mortality rates from this cause are extremely low.
<sup>c</sup> The model was restricted to ages 35 and older because mortality rates from this cause are exceptionally low below age 35.
<sup>d</sup> The model was restricted to ages 55 and older because there are very few deaths from Alzheimer's disease below age 55.

The bulk of excess mortality at the oldest ages resulted from COVID-19 (i.e., 80% or higher above age 65; Tables 1 and 2), but COVID-19 accounted for only a small fraction of the excess deaths at ages 15-24 (7% for males, 18% for females) and less than 35% of excess deaths at ages 25-44. Among males, 71% of the 59,271 non-COVID excess deaths occurred at working ages (25-64), but those ages accounted for only 36% of the 37,996 non-COVID excess deaths in females. Non-COVID excess mortality was more skewed towards older ages for females: ages 75 and older accounted for 44% of non-COVID excess deaths in females versus 11% in males.

The cause-specific models imply that, in males, the largest share of non-COVID excess deaths resulted from external causes (*N*=17,341), nearly 80% of which occurred at working ages (Table 1). For males, the relative increase in mortality from external causes was largest at ages 15-24 (Figure 3A), but the absolute number of excess deaths from external causes was highest at ages 25-34 (Figure 3B). In females, external causes contributed fewer excess deaths (*N*=3,536; Table 2) and both relative and absolute excess mortality from external causes were highest at ages 25-44 (eFigure 1).

**Figure 3.**
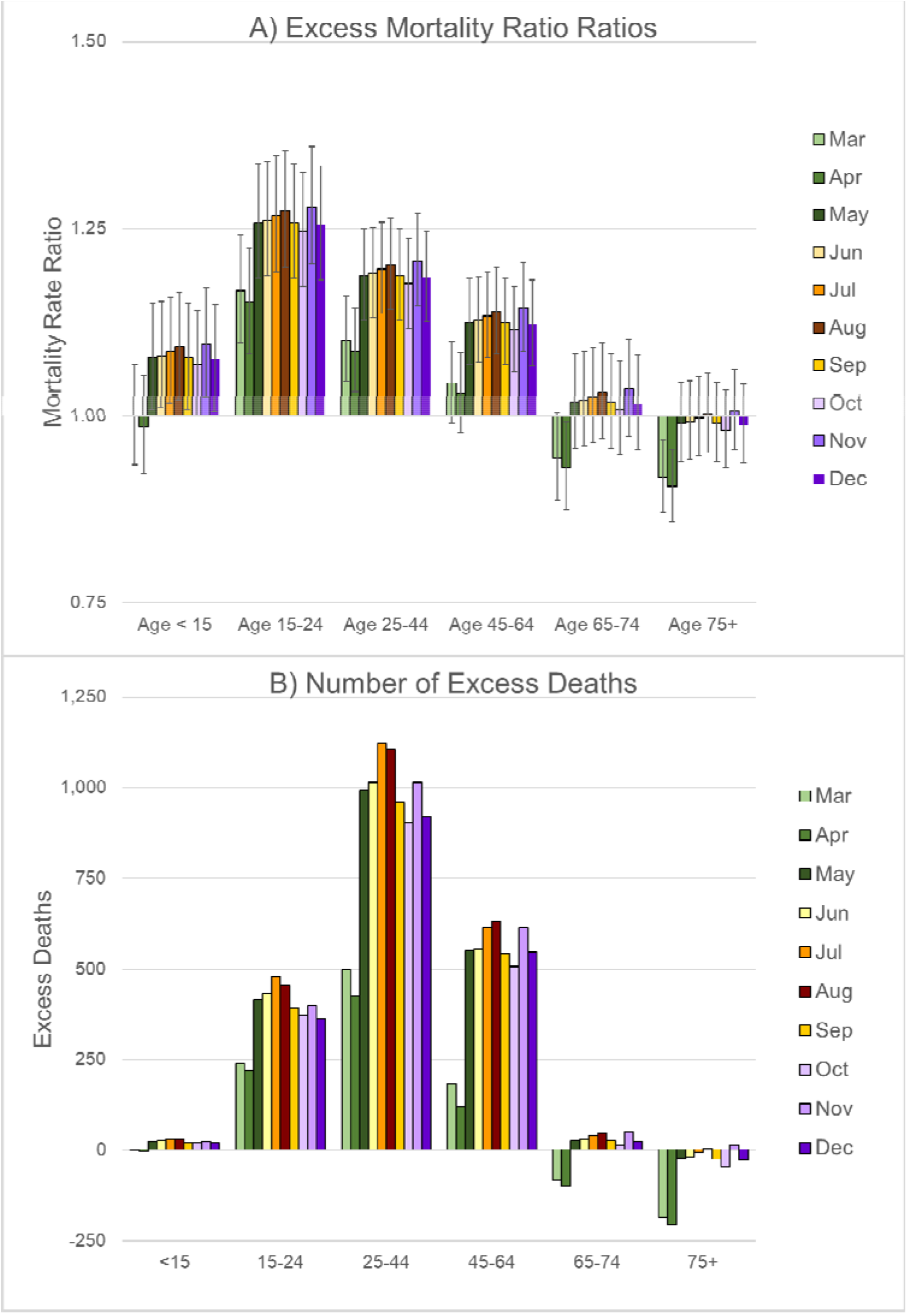
Excess Mortality from External Causes by Age Group During March-December 2020, US, Males: a) Rate Ratios; B) Number of Excess Deaths. Note: These estimates are based on the regression models shown in eTable 2. The bars on the graph of excess mortality rate ratios represent the 95% confidence interval.

Heart disease was the largest contributor of non-COVID excess deaths among females (*N*=10,566; Table 2) and the second biggest contributor among males (*N*=15,943; Table 1). The majority of excess deaths from heart disease occurred at working ages among males, but above age 65 in females.

Diabetes also accounted for a substantial number of excess deaths in both sexes (*N*=7,292 for males; *N*=6,433 for females). The excess mortality rate ratios for diabetes were highest at ages 25-44, but only slightly lower above age 45 (eFigures 2B & 3B). However, because pre-pandemic rates of diabetes mortality were much higher at older ages, the absolute number of excess deaths was much higher above age 45. Most of the excess deaths from diabetes occurred above age 65 (63% for males, 78% for females) and more than 92% were above age 45.

Alzheimer’s disease contributed a sizeable number of excess deaths among women (*N*=7,716), but less so in men (*N*=2,046). The vast majority of those deaths occurred above age 75 (87% for men, 91% for women).

Cerebrovascular disease also accounted for a notable share of excess deaths in both sexes. Most of those deaths occurred above age 65 (55% for men, 75% for women).

During March and April 2020, the excess mortality rate ratios for influenza and pneumonia were elevated at all ages (eFigures 2E & 3E), but mortality rates were significantly lower than expected in October-December 2020 among those aged 75 and older, especially women. When aggregated across the entire pandemic period (March-December 2020), influenza and pneumonia contributed 2,441 excess deaths in males, but no excess deaths in females because excess deaths in January-February were offset by lower than expected mortality later in the year.

There was little evidence of excess mortality from other respiratory diseases (eFigures 2F & 3F) or cancer (eFigures 2G & 3G). In fact, at the oldest ages, there were fewer deaths than expected for these two groups of causes (Tables 1 & 2).

### Sensitivity of the Results to Specification of the Pre-Pandemic Time Trend

For all-cause mortality (eFigure 4) and most of the cause-specific models (eFigure 5), the estimated number of excess deaths were similar across various specifications of the pre-pandemic time trend, although Model 1—which is based on the full time series with a linear pre-pandemic time trend—yielded the highest estimates for all causes, heart disease, cerebrovascular disease, diabetes, and external causes. For all of those outcomes, the estimates from Model 2 match those in Tables 1 and 2.

The causes for which the estimated number of excess deaths were the most sensitive to specification of the pre-pandemic time trend were heart disease, external causes, and cerebrovascular disease. More detailed analyses suggested that Model 1 probably over-estimated excess mortality, whereas Model 3 may have under-estimated excess mortality from these causes (see eText 4 for details). Among women, there was also considerable variation in the estimates of excess mortality from Alzheimer’s disease; Model 2 (which corresponds to the estimates provided in Table 2) had the best predictive accuracy for 2019, but also yielded the highest estimates of excess mortality from Alzheimer’s (eFigure 5).

The only other cause for which the main results indicated a substantial number of excess deaths was diabetes. The estimates of excess diabetes mortality presented in Tables 1 and 2 (i.e., Model 2 in eFigure 5) are somewhere in between the lowest and the highest values.

### Analysis of Sub-Categories of Mortality from External Causes

Unfortunately, monthly death counts by sex and age group are not yet available for sub-categories within external causes. Nonetheless, it was possible to analyze crude monthly death rates for drug overdoses, homicides, suicides, motor vehicle accidents, and unintentional injuries as a whole (see eText 5 for more details).^20^ Drug overdoses exhibited the highest excess mortality (peaking 49% higher than expected in May; RR=1.49, 95% CI=1.27-1.74) followed by homicides (peaking 43% higher than expected in October; RR=1.43, 95% CI=1.17-1.75). During March-May 2020, there was no evidence of significant excess mortality from motor vehicle accidents; in fact, rates were 16% *lower* than expected in April (RR=0.84, 95% CI=0.72-0.97). However, mortality from motor vehicle accidents was significantly higher than expected in June and August-December 2020, with the excess mortality rate ratio peaking at 1.20 (95% CI=1.03-1.39) in September 2020. As demonstrated in earlier studies, suicide mortality was significantly lower than expected throughout March-December 2020, with the biggest decline in April (−19%, HR=0.81, 95% CI=0.76-0.86).

## DISCUSSION

It is not surprising that heart disease, external causes, diabetes, Alzheimer’s disease, and cerebrovascular disease were major contributors of excess deaths because they were among the most common causes of death in the US during 2019.^21^ Even a small relative increase in mortality from these causes would generate lots of deaths because mortality rates were high even prior to the pandemic.

Diabetes exhibited a substantial relative increase in mortality across all ages 25 and older that persisted throughout April-December throughout 2020. To a lesser extent, excess mortality rate ratios were also elevated for heart disease, particularly at ages 25-74. Delayed health care for fear of infection, diversion of health care resources, and shortages of staff and equipment could have contributed to excess mortality from chronic diseases such as diabetes and heart disease. The social and economic consequences of the pandemic were also likely to pose a special challenge for people with chronic diseases that require careful daily management. Economic distress, disruptions of social life and support networks, and other stressors induced by the pandemic may have further hampered the management of chronic conditions. For diabetics, a lapse in glucose control or the temptation to ration insulin because of financial difficulty or loss of employer-sponsored health insurance, could have deadly consequences.

Although the results showed no evidence of excess cancer mortality, there could be a lagged effect. Delays in standard screening and reluctance to visit a doctor for a suspected problem may have reduced early detection and could later manifest as rising cancer mortality.

Misclassification of COVID-19 deaths could account for some excess mortality. For example, during March-April 2020, influenza and pneumonia exhibited the highest excess mortality in relative terms (eFigure 2E & 3E) with rate ratios as high as 1.75 among males aged 65-74 in March 2020, but this category is particularly likely to include misclassified COVID-19 deaths, especially early in the pandemic when testing for SARS-CoV-2 was limited. While misclassification of COVID-19 deaths could account for some of the excess mortality from influenza/pneumonia, circulatory diseases, and other natural causes, it is unlikely to explain the increase in mortality from external causes. Instead, excess mortality from injury-related causes was likely to be an indirect result of economic distress, the disruption of normal life, and heightened uncertainty. Analyses of crude death rates within sub-categories of external causes suggest that the increase in external mortality may have been driven primarily by drug-related mortality. While the pandemic may have exacerbated the drug epidemic, provisional data suggest that drug overdoses rose every month after February 2019, although the pace increased in the fall of 2019 and even more sharply after March 2020.^22^ Alcohol-related mortality may have also increased during the pandemic, but data are not yet available to answer that question. Early in the pandemic, many experts predicted that suicides would increase, yet the evidence suggests that suicide rates declined during April-December 2020.

### Limitations

Any estimate of excess mortality relies on the accuracy of hypothetical expected mortality (in the absence of the event of interest). Unfortunately, it is impossible to verify the accuracy of those estimates. Some of the apparent excess mortality could merely be a result of statistical noise. For example, the confidence intervals around the estimated rate ratios for excess Alzheimer’s disease mortality among men (eFigure 2C) were so wide that, in most cases, we cannot reject the null hypothesis (i.e., there was no excess mortality).

It is also important to acknowledge that one should not assume that all the estimated excess mortality is attributable to the pandemic. For example, if the model does not adequately account for the pre-pandemic increases in drug overdoses or if other factors unrelated to the pandemic contributed to increased drug mortality after March 2020, then the model would over-estimate excess drug overdoses associated with the pandemic.

These analyses are based on provisional death counts for 2020, which may be under-estimated, particularly during Wave 3. Because of reporting lags, some of the deaths for December are likely to be missing from the provisional data—which were last updated on July 27, 2021—especially for injury-related deaths (e.g., only 93% of drug overdoses are reported within 26 weeks after the death).^15^

Problems of misclassification are likely to plague the cause-specific mortality data. Over the course of the pandemic, as the availability of testing increased and medical examiners became more experienced identifying COVID-19, cause-of-death reporting undoubtedly improved. Thus, part of the decline in apparent excess mortality as the pandemic progressed could be a statistical artifact of more accurate reporting of COVID-19 deaths.

Data are not yet available to examine age and sex variation in excess mortality from more detailed causes such as drug- and alcohol-related mortality, suicide, and homicide. Once county-level data become available, this analysis will be extended to explore geographic variation in the impact of the pandemic on sub-categories of non-COVID mortality. There is already ample evidence that socioeconomically-disadvantaged communities were harder hit by COVID-19 than more advantaged communities.^5,23–25^ It seems likely that the pandemic further exacerbated socioeconomic disparities in drug- and alcohol-related mortality. Similarly, it will be interesting to investigate whether the decline in suicide benefited all communities equally. Well-off communities may have been able to mount a more effective response that benefited all residents, whereas more disadvantaged communities experienced further social disintegration.

Similarly, the data required to incorporate racial/ethnic variation into the model are not yet available. NCHS recently published provisional monthly death counts for selected causes of death by age, sex, and race/ethnicity,^14^ but that file uses a different racial classification than the bridged-race mortality data for 1999-2019.^13^ Inconsistency in racial classification could bias the results. It may not be possible to incorporate race/ethnicity into the current model until the final bridged-race death counts for 2020 are released.

### Policy Implications

The COVID-19 pandemic has exposed the gaping holes in America’s safety net. The most obvious implication of the results presented here is that disparities in access to high-quality healthcare exacerbate inequality in the US. The problem is not only lack of universal coverage. The US needs a healthcare system that rewards keeping patients healthy, rather than incentivizing greater use of unnecessary tests, procedures, and medication. As Case and Deaton^26^ point out, “The American health care industry is not good at promoting health, but it excels at taking money from all of us for its benefit. It is an engine of inequality.” An evidence-based, cost-effective health care system could help address some of the excess mortality highlighted here: better management of chronic health conditions could reduce morbidity and mortality from diabetes and heart disease; greater access to prompt acute care could minimize the impact of strokes; treatment for substance abuse and mental health disorders may help mitigate the fallout from the drug epidemic; and better options for relieving chronic pain could enhance quality of life and reduce the number of Americans who succumb to drug addiction.

Nonetheless, improving health care would only treat the symptoms of the underlying factors that leave some Americans vulnerable to chronic pain and illness, substance abuse, mental disorders, trauma, and cognitive impairment. Although this study did not highlight the socioeconomic, geographic, and racial/ethnic disparities in excess mortality, other research has exposed huge inequalities in the repercussions of the pandemic. Some of us have remained relatively unscathed by the pandemic, while others suffered heavy consequences to their health, financial wellbeing, and social environment. Policies need to address the cluster of social and economic factors that jeopardize the health and wellbeing of many Americans. Adverse outcomes and their underlying causes are intertwined and maintained by the intersection of socioeconomic stratification and structural racism. Policies that attempt to address the problems individually are not likely to be effective. For more than a year and a half, disadvantaged Americans have borne the brunt of the impact of the COVID-19 pandemic,^5,23–25^ but drug overdose mortality has been rising exponentially for at least 38 years.^27^ Even if it were possible to vaccinate everyone and eradicate SARS-CoV-2, that will not solve the ongoing drug epidemic in the US. There is no forthcoming vaccine for addiction. Whether we consider the current pandemic, the longstanding drug epidemic, rising prevalence of pain, declines in physical function, deteriorating mental health, or rising midlife mortality, the evidence suggests that disadvantaged Americans have suffered the most. .^23,28–35^

### Conclusion

Overall, most non-COVID excess deaths among males occurred at working ages (71%), whereas those ages accounted for only 36% of non-COVID excess deaths among females. Undoubtedly, the oldest Americans suffered the brunt of COVID-19 mortality, but working-age Americans, particularly men, suffered substantial increases in mortality from other causes of deaths during 2020. The number of excess deaths at younger ages may seem small compared with COVID-19 mortality at the oldest ages, but early deaths have a disproportionate effect on overall life expectancy. The US already had a midlife mortality crisis; the pandemic has deepened that crisis.

## Data Availability

All of the data used in this analysis are publicly available from the National Center for Health Statistics and the U.S. Census Bureau.

## ACKNOWLEDGMENTS

Special thanks to Maxine Weinstein, Magali Barbieri, Samuel H. Preston, Theresa Andrasfay, Andrew Stokes, and Noreen Goldman for their comments on early drafts of this manuscript. This research was supported by grants from the National Institute on Aging (R03 AG058110 and center grant P30 AG012839) and the Society of Actuaries.

## Footnotes

* Unless otherwise indicated, “excess mortality” includes deaths from all causes including COVID-19.

