## Supplementary Material for "THE US MIDLIFE MORTALITY CRISIS CONTINUES: EXCESS CAUSE-SPECIFIC MORTALITY DURING 2020"

### eText1. Data

The final death counts for 1999-2019^1^[From CDC Wonder for 1999-2019 (~Box\My Data\NCHS\CDC Wonder\Mortality Underlying COD\Monthly-Deaths-by-AgeGroup-and-Selected COD-1999-2019.xlsx and ~\Monthly-Deaths-by-AgeGroup-All Causes-1999-2019.xlsx)] include some deaths of unknown age. The maximum was 11 deaths of unknown age from external causes in November 1999, which represented less than 0.2% of all deaths from external causes in that month. Within each group defined by calendar year, month, sex, age, and the selected cause of death categories (see Measures section), we redistributed those deaths proportionately based on the distribution of deaths where age was known.

The data for 2020 remain provisional.^2^[Provisional estimates for Jan 2020 – June 2021 (~\Provisional\Deaths-by-COD, Age, Sex, and Race 2019-01-thru-2021-06.xlsx)]

### eText2. Cause of Death Groups

The cause-specific models were fit separately for the following 8 groups based on the ICD-10 codes for the underlying cause of death:^[1]^

1) Influenza and pneumonia (ICD-10 codes J09-J18);
2) Other respiratory diseases excluding COVID-19 (J00-J06,J30-J47,J67,J70-J98);
3) Heart diseases (I00-I09,I11,I13,I20-I51);
4) Cerebrovascular diseases (I60-I69);
5) Cancer (C00-C97);
6) Alzheimer’s disease (G30);
7) Diabetes mellitus (E10-E14); and
8) External causes (derived as deaths from all causes minus deaths from natural causes).^[2]^

Excess deaths for the residual category (i.e., all remaining causes of death) can be derived by subtracting the sum of estimated excess deaths across these 8 groups of causes from the estimated number of excess deaths from all-causes. When computing non-COVID mortality, we subtract the number of deaths where COVID-19 was reported to be the underlying cause.

### eText 3. Modeling Strategy

Monthly mortality rates by sex, age group, and selected cause of death were modeled separately by sex using negative binomial regression, which allows for an over-dispersed Poisson distribution (i.e., this model is appropriate for outcome variables generated by either a Poisson or a Poisson-like distribution in which variation is greater than that of a true Poisson). For each sex, the outcome variable was the number of deaths from a specified cause $(\boldsymbol{D}^{\boldsymbol{C}})$for a given month-year-age group, but the model controlled for the number of person-years of exposure $(\boldsymbol{E}\text{)}\boldsymbol{:}$

$\text{Ln} \boldsymbol{D}^{\text{C}}\boldsymbol{=}{\text{α}\boldsymbol{+}\boldsymbol{\beta}}_{\mathbf{a}}\boldsymbol{X}_{\mathbf{a}}\boldsymbol{+}\boldsymbol{\beta}_{\mathbf{m}}\boldsymbol{X}_{\mathbf{m}}\boldsymbol{+}\boldsymbol{\beta}_{\mathbf{ma}}\left( \boldsymbol{X}_{\mathbf{m}}\boldsymbol{\times}\boldsymbol{X}_{\text{a2}} \right)$

$$\boldsymbol{+}\boldsymbol{\beta}_{\mathbf{t}}\mathbf{T}\boldsymbol{+}\boldsymbol{\beta}_{\mathbf{t2}}\mathbf{T}^{\boldsymbol{2}}\boldsymbol{+}{\boldsymbol{\beta}_{\mathbf{t3}}\mathbf{T}^{\boldsymbol{3}}\boldsymbol{+}\boldsymbol{\beta}}_{\mathbf{ta}}\boldsymbol{(T \times}\boldsymbol{X}_{\mathbf{a}}\boldsymbol{)+} \boldsymbol{\beta}_{\mathbf{t2a}}\mathbf{(T}^{\boldsymbol{2}}\boldsymbol{\times}\boldsymbol{X}_{\mathbf{a}}\boldsymbol{)+} \boldsymbol{\beta}_{\mathbf{t3a}}\mathbf{(T}^{\boldsymbol{3}}\boldsymbol{\times}\boldsymbol{X}_{\mathbf{a}}\boldsymbol{)}$$

$\boldsymbol{+}\boldsymbol{\beta}_{\mathbf{m}}^{\boldsymbol{2020}}\mathbf{X}_{\mathbf{m}}^{\boldsymbol{2020}}\boldsymbol{+}\boldsymbol{\beta}_{\mathbf{P}}\left( \mathbf{P}\boldsymbol{\times}\boldsymbol{X}_{\mathbf{a3}} \right)\boldsymbol{+}\text{ln}\boldsymbol{(}\text{E}\boldsymbol{)+\epsilon}$.	(1)

Thus, the model can be rewritten with the logged death rate $\left( \ln\boldsymbol{M}^{\boldsymbol{C}} \right)$, where $\boldsymbol{M}^{\boldsymbol{C}}$ represents deaths $(\boldsymbol{D}^{\boldsymbol{C}})$ divided by exposure $(\text{E}\text{)}$, on the left-hand side:

$\text{Ln} \boldsymbol{M}^{\boldsymbol{C}}\boldsymbol{=}{\text{α}\boldsymbol{+}\boldsymbol{\beta}}_{\mathbf{a}}\boldsymbol{X}_{\mathbf{a}}\boldsymbol{+}\boldsymbol{\beta}_{\mathbf{m}}\boldsymbol{X}_{\mathbf{m}}\boldsymbol{+}\boldsymbol{\beta}_{\mathbf{ma}}\left( \boldsymbol{X}_{\mathbf{m}}\boldsymbol{\times}\boldsymbol{X}_{\text{a2}} \right)$

$$\boldsymbol{+}\boldsymbol{\beta}_{\mathbf{t}}\boldsymbol{T+}\boldsymbol{\beta}_{\mathbf{t2}}\boldsymbol{T}^{\boldsymbol{2}}\boldsymbol{+}{\boldsymbol{\beta}_{\mathbf{t3}}\boldsymbol{T}^{\boldsymbol{3}}\boldsymbol{+}\boldsymbol{\beta}}_{\mathbf{ta}}\mathbf{(}\boldsymbol{T}\boldsymbol{\times}\boldsymbol{X}_{\mathbf{a}}\boldsymbol{)+} \boldsymbol{\beta}_{\mathbf{t2a}}{\mathbf{(}\boldsymbol{T}}^{\boldsymbol{2}}\boldsymbol{\times}\boldsymbol{X}_{\mathbf{a}}\boldsymbol{)+} \boldsymbol{\beta}_{\mathbf{t3a}}{\mathbf{(}\boldsymbol{T}}^{\boldsymbol{3}}\boldsymbol{\times}\boldsymbol{X}_{\mathbf{a}}\boldsymbol{)}$$

$\boldsymbol{+}\boldsymbol{\beta}_{\mathbf{m}}^{\boldsymbol{2020}}\mathbf{X}_{\mathbf{m}}^{\boldsymbol{2020}}\boldsymbol{+}\boldsymbol{\beta}_{\mathbf{P}}\boldsymbol{(P\times}\boldsymbol{X}_{\mathbf{a3}}\boldsymbol{)}$ $\boldsymbol{+\epsilon}$.	(2)

The first part of the model was designed to capture levels and trends in background mortality prior to the pandemic: $\boldsymbol{X}_{\mathbf{a}}$ represents a set of dummy variables for each age group (i.e., 0-4, 5-14,…74-84, 85+) to account for the age pattern of mortality; $\boldsymbol{X}_{\mathbf{m}}$ is a set of dummy variables for each month to capture the seasonality of mortality; and $\boldsymbol{T}$ denotes calendar year. In preliminary analyses, data for 1999-2018 (20-year period) were used to test several specifications for the pre-pandemic time trend (i.e. linear, quadratic, cubic, dummy for each calendar year). The coefficients from each model were used to predict the number of deaths in 2019. The resulting residuals were then used to compute the root mean squared error (RMSE) to determine the specification that best predicted mortality in 2019. Since it is impossible to know how many deaths would have occurred in 2020 in the absence of a pandemic, the model that best predicts mortality in 2019 is likely to provide the best estimate of expected mortality in 2020.

There was no significant time trend for influenza/pneumonia mortality; the linear specification exhibited the best predictive accuracy for 2019 (i.e., lowest RMSE). For several other outcomes (i.e., all-cause, cardiovascular disease, Alzheimer’s disease, diabetes, external causes, ill-defined causes, and the residual category), the quadratic specification yielded the lowest RMSE. For other respiratory deaths and cancer, the cubic specification had the best predictive accuracy for 2019. All models also included interactions between the time trend parameters and age because mortality decline has been slower or even reversed among working-age Americans during recent years compared with older Americans, especially those aged 80 and older.^4^

To evaluate the sensitivity of the results to the length of the time series used to fit the model, the models were refit using data only for 10-year (2009-18) and 5-year (2014-18) periods with a linear time trend. [For such short time series, there was not enough information to identify a quadratic or cubic time trend interacted with age; in many cases, the model would not even converge.] For three outcomes (i.e., heart disease, cerebrovascular disease, and diabetes), the model based on 2014-18 predicted deaths in 2019 somewhat better than the model based on 1999-2018 with a quadratic time trend. The model based on 2009-18 never yielded the lowest RSME (perhaps because the linear time trend could not capture potential non-linearities, but the series was too short to allow for a non-linear time trend).

Prior work has demonstrated that seasonality can vary by age; for example, all-cause mortality below age 5 and above age 35 peaks in January or February, whereas mortality between age 5 and 35 peaks in July-September.^5^[see Fig 11 of Parks et al. 2018.] However, that study found little evidence that seasonality varies by age for some specific causes (e.g., cardiorespiratory). Thus, we tested an interaction between month $\boldsymbol{(X}_{\mathbf{m}}\boldsymbol{)}$and a set of dummy variables for three age groups (i.e., <5, 5-34, 35+, represented by $\boldsymbol{X}_{\mathbf{a2}}\boldsymbol{)}$. These interactions improved model fit (based on the Bayesian information criterion) only for all-cause mortality and external causes. Therefore, we included the month-by-age group interactions in the models for only those outcomes.

The second part of the model was designed to determine the extent to which mortality in 2020 differed from the pre-pandemic time trend. The model included an additional set of dummy variables for each month of 2020 ($\mathbf{X}_{\mathbf{m}}^{\boldsymbol{2020}})$. To test whether excess mortality during the pandemic varied by age, the model also included interactions between a dichotomous indicator ($\boldsymbol{P}\mathbf{),}$ which denoted the period after COVID-19 was declared a pandemic (March 2020-), and a set of dummy variables for broad age groups (i.e., <15, 15-24, 25-44, 45-64, 65-74, 75+, represented by $\boldsymbol{X}_{\mathbf{a3}}\boldsymbol{)}$. For this interaction, several of the age groups for which the risk of COVID-19 mortality was likely to be similar were combined. The pandemic-by-age interactions improved model fit for all outcomes.

In addition, an auxiliary model was estimated to evaluate whether excess mortality varied by both age and wave of the pandemic (i.e., Wave 1: March-June; Wave 2: July-September; Wave 3: October-December).^[3]^ As shown in Equation (3), this model added interactions between a dummy variable for each wave ($\boldsymbol{W}_{\boldsymbol{2}}$ for Wave 2 and $\boldsymbol{W}_{\boldsymbol{3}}$ for Wave 3, with Wave 1 serving as the reference group) and the set of dichotomous variables for the broad age groups $(\boldsymbol{X}_{\mathbf{a2}}\boldsymbol{)}$. The specification of the time trend was the same as in the previous set of models, although Equation (3) shows the cubic specification.

$\text{Ln} \boldsymbol{M}^{\boldsymbol{C}}\boldsymbol{=}{\text{α}\boldsymbol{+}\text{β}}_{\text{a}}\boldsymbol{X}_{\mathbf{a}}\boldsymbol{+}\boldsymbol{\beta}_{\mathbf{m}}\boldsymbol{X}_{\mathbf{m}}\boldsymbol{+}\boldsymbol{\beta}_{\mathbf{ma}}\left( \boldsymbol{X}_{\mathbf{m}}\boldsymbol{\times}\boldsymbol{X}_{\text{a2}} \right)$

$$\boldsymbol{+}\boldsymbol{\beta}_{\mathbf{t}}\boldsymbol{T+}\boldsymbol{\beta}_{\mathbf{t2}}\boldsymbol{T}^{\boldsymbol{2}}\boldsymbol{+}{\boldsymbol{\beta}_{\mathbf{t3}}\boldsymbol{T}^{\boldsymbol{3}}\boldsymbol{+}\boldsymbol{\beta}}_{\mathbf{ta}}\mathbf{(}\boldsymbol{T}\boldsymbol{\times}\boldsymbol{X}_{\mathbf{a}}\boldsymbol{)+} \boldsymbol{\beta}_{\mathbf{t2a}}{\mathbf{(}\boldsymbol{T}}^{\boldsymbol{2}}\boldsymbol{\times}\boldsymbol{X}_{\mathbf{a}}\boldsymbol{)+} \boldsymbol{\beta}_{\mathbf{t3a}}{\mathbf{(}\boldsymbol{T}}^{\boldsymbol{3}}\boldsymbol{\times}\boldsymbol{X}_{\mathbf{a}}\boldsymbol{)}$$

$\boldsymbol{+}\boldsymbol{\beta}_{\mathbf{m}}^{\boldsymbol{2020}}\mathbf{X}_{\mathbf{m}}^{\boldsymbol{2020}}\boldsymbol{+}\boldsymbol{\beta}_{\mathbf{P}}\boldsymbol{(P\times}\boldsymbol{X}_{\mathbf{a3}}\boldsymbol{)} \boldsymbol{+}\boldsymbol{\beta}_{\mathbf{W2}}\left( \boldsymbol{W}_{\boldsymbol{2}}\boldsymbol{\times}\boldsymbol{X}_{\mathbf{a3}} \right)\boldsymbol{+} \boldsymbol{\beta}_{\mathbf{W3}}\left( \boldsymbol{W}_{\boldsymbol{3}}\boldsymbol{\times}\boldsymbol{X}_{\mathbf{a3}} \right)\boldsymbol{+\epsilon}$.	(3)

### eText 4. Sensitivity to Model Specification

eFigures 4 and 5 demonstrate the sensitivity of the results five different specifications for the pre-pandemic time trend. The first three specifications were fit to the full data series (1999-). The only difference was that Model 1 assumed the pre-pandemic time trend was linear, whereas Model 2 used a quadratic specification and Model 3 comprised a cubic specification.

Nepomuceno et al.^7^ showed that the length of the reference period used to fit the model also affects the estimates of excess mortality. Thus, the last two models were fit to a shorter data series (2009- for Model 4; 2014- for Model 5). Both used a specification similar to Model 1 (i.e., linear time trend) albeit more simplified because the shorter time series did not provide sufficient variation to identify all the parameters in a more complicated specification (see footnotes to eTable 4 for more details).

#### 4.1 All-Cause Mortality

For all-cause mortality (eFigure 4), Model 1 (1999-, linear time trend) yielded the highest estimates of excess mortality (*N*=315,622 excess deaths for males, *N*=229,842 for females), whereas Model 3 (1999-, cubic time trend) produced the lowest estimates (*N*=223,701 for males, *N*=177,015 for females). The estimates from Model 2 (1999-, quadratic time trend), Model 4 (2009‑, linear time trend), and Model 5 (2014-, linear time trend) were similar (252,199-258,929 for males, 194,884-1999,507 for females). In terms of overall predictive accuracy, Model 3 performed the best (i.e., lowest RMSE), while Model 1 fared the worst (eTable 4). For 2019 (i.e., the period immediately prior to the pandemic), Model 2 yielded the best predictive accuracy, but Model 1 was still the worst. Examination of the residuals for 2019 revealed that Model 1 generally under-estimated deaths, whereas Model 3 typically over-estimated deaths.

An assessment of the variation in estimates of excess mortality by sex and age group showed that the biggest differences across models were among men aged 65-74 (i.e., estimates of excess mortality ranged from 47,252 for Model 3 to 84,631 for Model 1, a difference of nearly 38,000 deaths). eFigure 6 shows the observed versus expected monthly death counts for men aged 65-74 during 2019-2020. Prior to March 2020, there should be a close match between the observed and expected counts; during the pandemic period (March 2020-), the difference between the observed and expected represents the estimate of excess mortality. The plot indicates that Model 1 (1999- with a linear time trend) consistently under-estimated mortality during 2019. Thus, it is reasonable to conclude that this model probably also under-estimated expected mortality during 2020-21, which would lead to an over-estimate of excess mortality.

In contrast, the other four models appeared to predict the observed death counts in 2019 rather well. Although the overall differences in predictive accuracy between those models were small, among men aged 65-74, the fitted counts from Model 3 (1999-, cubic specific for the time trend) were consistently the highest (eFigure 6). Indeed, the expected counts based on Model 3 were higher than the observed death counts for 12 of the 14 months between January 2019 and February 2020. Thus, it seems likely that Model 3 also over-estimated expected mortality during the pandemic and thus, under-estimated excess mortality.

As noted in eText 1, when the models were fit only to the data prior to 2019, Model 2 (1999- with a linear time trend) predicted all-cause mortality in 2019 the best. Therefore, those are the estimates presented in the main analyses (Tables 1 & 2). In males, the estimated number of excess deaths based on Model 2 was the second lowest across all five models whereas it was the median estimate for females (eFigure 4).

#### 4.2 Cause-Specific Mortality

eFigure 5 shows the variation in the estimates of excess mortality by cause of death. For heart disease, cerebrovascular disease, diabetes, and external causes, Model 1 (1999-, linear time trend) yielded the highest estimates, Model 3 (cubic, 1999-) produced the smallest estimates, and the estimates from Models 2, 4, and 5 were generally similar. For most outcomes, Model 1 yielded the worst overall predictive accuracy; the only exceptions were influenza/pneumonia, cancer, and Alzheimer’s disease (eTable 4). An examination of the residuals for 2019 showed that Model 1 generally under-estimated deaths from heart disease and cerebrovascular disease in both sexes and under-estimated deaths from external causes among males, whereas Model 3 tended to over-estimated deaths from those same causes. In the case of Alzheimer’s disease, Model 3 and to a lesser extent models 1, 4, and 5 tended to over‑estimate deaths in 2019; Model 2 yielded the smallest residuals for Alzheimer’s disease in 2019.

The greatest variation in the estimated number of excess deaths was from heart disease among males (e.g., Model 1 indicated 46,810 excess deaths, whereas Model 3 suggests excess mortality was only 7,614; the other estimates ranged from 15,943 to 22,162). A closer look at the estimates by age group showed that the biggest differences were among women aged 85 and older (i.e., estimated excess mortality ranged from -4,529 for Model 3 to 14,542 for Model 1). Thus, eFigure 7 graphs the observed versus expected heart disease deaths among women aged 85 and older during 2019-2020. Among this subgroup, Model 1 consistently under‑estimated deaths in 2019, whereas Model 3 generally over-estimated mortality. Thus, during the pandemic period, it seems likely that Model 1 over-estimated excess mortality and Model 3 under-estimated excess mortality. The highest number of heart disease deaths among this subgroup occurred in April 2020; eFigure 8 plots the observed versus expected number of heart disease deaths during April across the entire period since 1999. Again, this graph clearly shows that Model 1 did not fit well prior to the pandemic: the fitted values were consistently lower than the observed death counts for the 5-year period (2015-19) immediately prior to the pandemic, which suggests that Model 1 may over-estimate excess mortality during the pandemic. The other four models did better at predicting deaths in 2019 in this subgroup, but Model 3 implied that heart disease deaths would continue to increase between 2019 and 2020 (which would yield lower estimates of excess mortality), whereas Models 2, 4, and 5 implied that expected deaths in 2020 would be similar to or lower than in 2019 (which would yield higher estimates of excess mortality). The range of estimated total number of excess heart disease deaths (across all ages 15+) across those four models was 7,614 to 22,162 for males and -666 to 13,252 for females. The estimates presented in Tables 1 and 2 (which are based on Model 2) are higher than the estimates based on Model 3, but lower than Models 4 or 5.

There was also a lot of variability in the estimates of excess mortality from external causes, especially among males (eFigure 5). An examination of the estimates for males by age group revealed that the biggest differences were at ages 25-34. Among that subgroup (eFigure 9), Model 1 under-estimated the number of deaths during January – March 2019, fit rather well during April-August 2019, and greatly under-estimated deaths during September 2019 through February 2020. Thus, it seems likely that Model 1 continued to under-estimate expected mortality (in the absence of a pandemic) for the period since March 2020, which would imply that the estimates of excess mortality based on Model 1 were too high. On the other hand, Model 3 consistently over-estimated deaths during 2019. The models that most closely matched the observed death counts during the 6-month period prior to the pandemic (i.e., September 2019 – February 2020) were Models 2 and 4. During the pandemic, the highest number of deaths among men aged 25-34 occurred in August 2020. eFigure 10 plots the deaths during August from external causes among this subgroup for the entire period since 1999. Observed deaths from external causes among men in this age group were generally increasing from 1999 to 2019 (before the pandemic), but there was a sharp increase between 2015 and 2016, a decline in 2018, and then a rise back to 2017 levels in 2019. Thus, it is difficult to predict expected deaths from external causes in 2020 in the absence of a pandemic. Nonetheless, the preponderance of evidence suggests that Model 1 probably over-estimated excess mortality (because estimates of expected mortality were too low), whereas Model 3 probably under-estimated excess mortality. It seems likely that excess deaths probably somewhere within the estimates for Models 2, 4, and 5, which ranged between 11,825 and 17,341 for males. The estimates presented in Table 2 (based on Model 2) were the highest among those three models.

It may seem surprising that the estimates of excess mortality from Alzheimer’s disease (AD) are so low. There is no doubt that the absolute number of deaths from AD increased from 2019 to 2020.^8^ Yet, some increase in the death count would be expected simply as a result of population growth and aging of the population. The largest number of deaths from AD and the greatest variation in estimates of excess mortality from AD occurred among women ages 85 and older. Among this subgroup (eFigure 11), most of the models over-estimated the number of deaths from AD during the pre-pandemic period (January 2019-February 2020). Model 2 provided the best match between observed and fitted death counts during that period. Therefore, it seems fair to assume that Model 2 would provide the best estimate of expected mortality in the absence of a pandemic after March 2020. Those are the estimates presented in Table 2, but it is important to note that they were the highest estimates across all five models. If the estimates of expected AD mortality based on Model 2 were too low, then the estimates of excess mortality are over-estimated.

### eText 5. Analyses for Sub-Categories of External Causes

More detailed data for the following sub-categories of external causes are available through March 31, 2021,^3^ but those data are not disaggregated by sex or age group. The data for other causes of death are available through October 2, 2021, but deaths for subgroups of external causes (i.e., accidents, traffic-related fatalities, suicides, homicides, and drug overdoses) are available only through March 31, 2021 because the National Center for Health Statistics imposes a six-month lag on the release of those death counts in order to ensure the data are reasonably complete.^9,^^[4]^

The category for drug overdoses includes all drug overdoses (ICD-10: X40-X44, X60-X64, X85, Y10-14), regardless of intent; thus, it overlaps with homicides and suicides. Homicides include assault by drugs, medicaments, and biological substances (X85). Suicides include intentional drug overdoses (X60-X64). The broad category for unintentional injuries includes unintentional drug overdoses.

These models were similar to the model for external causes described in the main text, but because sex- and age-specific data are not yet available, it was impossible to evaluate the extent to which excess mortality for these causes varied by sex and age. Thus, a negative binomial model was used to regress the log of the monthly death rate for each sub-category of external causes $\left( \ln\boldsymbol{M}^{\boldsymbol{C}} \right)$ on the set of dummy variables for each month $\boldsymbol{(X}_{\mathbf{m}}$), which captures the seasonality of mortality; a quadratic specification for the historical time trend, where $\boldsymbol{T}$ denotes calendar year; and additional set of dummy variables for each month of 2020 ($\mathbf{X}_{\mathbf{m}}^{\boldsymbol{2020}})$ to determine the extent to which mortality in 2020 differed from the pre-pandemic time trend:

$\text{Ln} \boldsymbol{M}^{\boldsymbol{C}}\boldsymbol{=}\text{ α}\boldsymbol{+}\text{ }\boldsymbol{\beta}_{\mathbf{m}}\boldsymbol{X}_{\mathbf{m}}\boldsymbol{+}\boldsymbol{\beta}_{\mathbf{t}}\boldsymbol{T+}\boldsymbol{\beta}_{\mathbf{t2}}\boldsymbol{T}^{\boldsymbol{2}}\boldsymbol{+}\boldsymbol{\beta}_{\mathbf{m}}^{\boldsymbol{2020}}\boldsymbol{X}_{\mathbf{m}}^{\boldsymbol{2020}}$.	(4)

|  | **Males** | | **Females** | |
| --- | --- | --- | --- | --- |
|  | Coef | 95% CI | Coef | 95% CI |
| Pre-pandemic portion of 2020 | | | | |
| January 2020 | -0.01 | (-0.04 - 0.02) | -0.02 | (-0.05 - 0.01) |
| February 2020 | 0.03 | (-0.01 - 0.06) | 0.01 | (-0.02 - 0.04) |
| Pandemic period (Mar-Dec 2020) | | | | |
| March 2020 | 0.04** | (0.00 - 0.07) | 0.02 | (-0.01 - 0.06) |
| April 2020 | 0.23*** | (0.20 - 0.27) | 0.21*** | (0.17 - 0.24) |
| May 2020 | 0.16*** | (0.13 - 0.20) | 0.14*** | (0.10 - 0.17) |
| June 2020 | 0.13*** | (0.09 - 0.16) | 0.12*** | (0.09 - 0.16) |
| July 2020 | 0.19*** | (0.15 - 0.23) | 0.18*** | (0.15 - 0.22) |
| August 2020 | 0.17*** | (0.13 - 0.21) | 0.18*** | (0.15 - 0.22) |
| September 2020 | 0.14*** | (0.10 - 0.18) | 0.12*** | (0.09 - 0.16) |
| October 2020 | 0.13*** | (0.09 - 0.17) | 0.12*** | (0.08 - 0.15) |
| November 2020 | 0.20*** | (0.17 - 0.24) | 0.19*** | (0.16 - 0.22) |
| December 2020 | 0.27*** | (0.23 - 0.30) | 0.24*** | (0.21 - 0.27) |
| Interactions: Pandemic period (dummy for Mar-Dec 2020) x | | | | |
| Age < 15 | -0.20*** | (-0.24 - -0.16) | -0.23*** | (-0.27 - -0.19) |
| Age 15-24 | 0.05** | (0.01 - 0.09) | 0.00 | (-0.05 - 0.04) |
| Age 25-44 | 0.06*** | (0.03 - 0.09) | 0.03* | (-0.00 - 0.06) |
| Age 45-64 | 0.04*** | (0.01 - 0.08) | 0.00 | (-0.03 - 0.03) |
| Age 65-74 | 0.01 | (-0.03 - 0.05) | 0.01 | (-0.02 - 0.05) |
| Age 75+ | *(omitted)* |  | *(omitted)* |  |
| Alpha | 0.002 |  | 0.001 |  |
| Constant | 5.10*** | (5.08 - 5.13) | 4.90*** | (4.87 - 4.92) |
| Observations | 2,640 |  | 2,640 |  |

* p<0.05, ** p<0.01, *** p<0.001

Note: Both models also control for the main effects of age (<5,5-14,25-34,…75-84,85+), month, and calendar year (quadratic specification) and include the following interactions: age x (year, year squared); month x age group (<5, 5-34, 35+).

eTable 2. Coefficients (95% CI) from negative binomial regression models predicting mortality for selected causes of death in the US, 1999-2020, Males

|  | **Influenza/Pneumonia** | | **Other Respiratory** | | **Heart Disease** | | **Cerebrovascular** | |
| --- | --- | --- | --- | --- | --- | --- | --- | --- |
|  | Coef | 95% CI | Coef | 95% CI | Coef | 95% CI | Coef | 95% CI |
| Pre-pandemic portion of 2020 | | | | | | | | |
| January 2020 | 0.13* | (-0.01 - 0.27) | -0.04 | (-0.09 - 0.01) | -0.05*** | (-0.08 - -0.02) | -0.03 | (-0.07 - 0.02) |
| February 2020 | 0.24*** | (0.11 - 0.38) | -0.02 | (-0.07 - 0.03) | -0.02 | (-0.05 - 0.01) | -0.01 | (-0.06 - 0.03) |
| Pandemic period (Mar-Dec 2020) | | | | | | | | |
| March 2020 | 0.34*** | (0.20 - 0.49) | -0.03 | (-0.09 - 0.02) | -0.06*** | (-0.09 - -0.02) | -0.02 | (-0.07 - 0.03) |
| April 2020 | 0.34*** | (0.19 - 0.49) | 0.02 | (-0.03 - 0.08) | 0.05*** | (0.02 - 0.09) | 0.04* | (-0.01 - 0.09) |
| May 2020 | -0.03 | (-0.18 - 0.12) | -0.08*** | (-0.14 - -0.03) | 0.01 | (-0.03 - 0.04) | -0.02 | (-0.07 - 0.03) |
| June 2020 | -0.1 | (-0.25 - 0.06) | -0.07** | (-0.13 - -0.02) | 0.03 | (-0.01 - 0.06) | 0.03 | (-0.02 - 0.08) |
| July 2020 | 0 | (-0.15 - 0.16) | -0.01 | (-0.07 - 0.05) | 0.07*** | (0.03 - 0.10) | 0.05* | (-0.00 - 0.09) |
| August 2020 | 0.03 | (-0.12 - 0.18) | 0 | (-0.06 - 0.06) | 0.06*** | (0.03 - 0.10) | 0.05** | (0.01 - 0.10) |
| September 2020 | 0.01 | (-0.15 - 0.16) | -0.02 | (-0.07 - 0.04) | 0.03* | (-0.00 - 0.07) | 0.05* | (-0.00 - 0.10) |
| October 2020 | -0.15* | (-0.30 - 0.00) | -0.04 | (-0.09 - 0.02) | 0.02 | (-0.01 - 0.06) | 0.04* | (-0.01 - 0.09) |
| November 2020 | -0.12 | (-0.27 - 0.03) | -0.05* | (-0.11 - 0.00) | 0.02 | (-0.01 - 0.06) | 0.02 | (-0.03 - 0.06) |
| December 2020 | -0.15** | (-0.30 - -0.00) | -0.09*** | (-0.15 - -0.04) | 0.05*** | (0.01 - 0.08) | 0.04* | (-0.00 - 0.09) |
| Pandemic x |  |  |  |  |  |  |  |  |
| Age < 15 |  |  |  |  |  |  |  |  |
| Age 15-24 |  |  |  |  | -0.03 | (-0.14 - 0.08) |  |  |
| Age 25-44 | 0.11 | (-0.05 - 0.27) | 0.30*** | (0.20 - 0.39) | 0.16*** | (0.12 - 0.19) | 0.13*** | (-0.02 - 0.05) |
| Age 45-64 | 0.15*** | (0.04 - 0.26) | 0.09*** | (0.04 - 0.14) | 0.07*** | (0.05 - 0.10) | 0.09*** | (0.00 - 0.03) |
| Age 65-74 | 0.21*** | (0.08 - 0.34) | 0.02 | (-0.03 - 0.08) | 0.01 | (-0.03 - 0.04) | 0.03 | (-0.01 - 0.02) |
| Age 75+ | *(omitted)* |  | *(omitted)* |  | *(omitted)* |  | *(omitted)* |  |
| Alpha | 0.023 |  | 0.002 |  | 0.001 |  | 0.001 |  |
| Constant | 1.15*** | (1.09 - 1.21) | 0.36*** | (0.27 - 0.45) | 1.10*** | (1.05 - 1.15) | 0.36*** | (0.29 - 0.43) |
| Observations | 1,584 |  | 1,848 |  | 2,112 |  | 2,640 |  |

**eTable 2 (Continued)**

|  | **Cancer** | | **Alzheimer's Disease** | | **Diabetes Mellitus** | | **External Causes** | |
| --- | --- | --- | --- | --- | --- | --- | --- | --- |
|  | Coef | 95% CI | Coef | 95% CI | Coef | 95% CI | Coef | 95% CI |
| Pre-pandemic portion of 2020 | | | | | | | | |
| January 2020 | 0.01 | (-0.01 - 0.02) | -0.10** | (-0.18 - -0.02) | -0.04* | (-0.09 - 0.00) | 0.01 | (-0.04 - 0.05) |
| February 2020 | 0.03*** | (0.01 - 0.04) | -0.07 | (-0.15 - 0.02) | 0.03 | (-0.01 - 0.08) | 0.05** | (0.01 - 0.10) |
| Pandemic period (Mar-Dec 2020) | | | | | | | | |
| March 2020 | 0.00 | (-0.02 - 0.02) | 0.02 | (-0.06 - 0.11) | 0.04* | (-0.01 - 0.09) | -0.09*** | (-0.14 - -0.03) |
| April 2020 | 0.00 | (-0.02 - 0.01) | 0.14*** | (0.05 - 0.22) | 0.23*** | (0.19 - 0.28) | -0.10*** | (-0.15 - -0.05) |
| May 2020 | -0.02*** | (-0.04 - -0.01) | 0.05 | (-0.04 - 0.13) | 0.13*** | (0.08 - 0.18) | -0.01 | (-0.06 - 0.04) |
| June 2020 | -0.02** | (-0.04 - -0.00) | 0.06 | (-0.03 - 0.15) | 0.09*** | (0.04 - 0.14) | -0.01 | (-0.06 - 0.04) |
| July 2020 | 0.01 | (-0.01 - 0.02) | 0.12*** | (0.04 - 0.21) | 0.21*** | (0.16 - 0.25) | 0.00 | (-0.06 - 0.05) |
| August 2020 | 0.00 | (-0.01 - 0.02) | 0.07 | (-0.02 - 0.16) | 0.17*** | (0.12 - 0.22) | 0.00 | (-0.05 - 0.05) |
| September 2020 | 0.00 | (-0.02 - 0.02) | 0.04 | (-0.05 - 0.13) | 0.13*** | (0.08 - 0.18) | -0.01 | (-0.06 - 0.04) |
| October 2020 | -0.01 | (-0.03 - 0.01) | -0.02 | (-0.11 - 0.07) | 0.10*** | (0.05 - 0.15) | -0.02 | (-0.07 - 0.03) |
| November 2020 | 0.00 | (-0.01 - 0.02) | 0.05 | (-0.04 - 0.14) | 0.15*** | (0.11 - 0.20) | 0.01 | (-0.05 - 0.06) |
| December 2020 | -0.01 | (-0.02 - 0.01) | 0.06 | (-0.03 - 0.14) | 0.22*** | (0.17 - 0.27) | -0.01 | (-0.06 - 0.04) |
| January 2021 | 0.00 | (-0.02 - 0.02) | 0.02 | (-0.06 - 0.11) | 0.04* | (-0.01 - 0.09) | -0.09*** | (-0.14 - -0.03) |
| February 2021 | 0.00 | (-0.02 - 0.01) | 0.14*** | (0.05 - 0.22) | 0.23*** | (0.19 - 0.28) | -0.10*** | (-0.15 - -0.05) |
| Pandemic x |  |  |  |  |  |  |  |  |
| Age < 15 | -0.04 | (-0.16 - 0.07) |  |  |  |  | 0.08*** | (0.02 - 0.15) |
| Age 15-24 | 0.02 | (-0.08 - 0.12) |  |  |  |  | 0.24*** | (0.18 - 0.30) |
| Age 25-44 | 0.01 | (-0.03 - 0.05) |  |  | 0.13*** | (0.07 - 0.19) | 0.18*** | (0.13 - 0.23) |
| Age 45-64 | 0.02* | (-0.00 - 0.03) | -0.01 | (-0.13 - 0.11) | 0.02 | (-0.01 - 0.06) | 0.13*** | (0.08 - 0.17) |
| Age 65-74 | 0.01 | (-0.01 - 0.02) | 0.02 | (-0.05 - 0.10) | 0.00 | (-0.04 - 0.04) | 0.03 | (-0.03 - 0.09) |
| Age 75+ | *(omitted)* |  | *(omitted)* |  | *(omitted)* |  | *(omitted)* |  |
| Alpha | 0.000 |  | 0.004 |  | 0.001 |  | 0.003 |  |
| Constant | 0.90*** | (0.81 - 0.99) | 0.64*** | (0.56 - 0.72) | 0.57*** | (0.51 - 0.63) | 2.79*** | (2.74 - 2.84) |
| Observations | 2,640 |  | 1,056 |  | 1,848 |  | 2,640 |  |

* p<0.05, ** p<0.01, *** p<0.001

Note: All models also controlled for the main effects of age (<5,5-14,25-34,…75-84,85+), month, and calendar year (linear specification for influenza/pneumonia, cubic specification for other respiratory diseases and cancer; quadratic specification for the remaining outcomes). In addition, the models included interactions between the age groups and the time trend (i.e., linear, quadratic or cubic). The model for external causes also included interactions between month and broad age groups (i.e., <5, 5-34, 35+).

eTable 3. Coefficients (95% CI) from negative binomial regression models predicting mortality for selected causes of death in the US, 1999-2020, Females

|  | **Influenza/Pneumonia** | | **Other Respiratory** | | **Heart Disease** | | **Cerebrovascular** | |
| --- | --- | --- | --- | --- | --- | --- | --- | --- |
|  | Coef | 95% CI | Coef | 95% CI | Coef | 95% CI | Coef | 95% CI |
| Pre-pandemic portion of 2020 | | | | | | | | |
| January 2020 | 0.15* | (-0.00 - 0.31) | -0.06** | (-0.12 - -0.01) | -0.06*** | (-0.09 - -0.03) | -0.04 | (-0.08 - 0.01) |
| February 2020 | 0.23*** | (0.07 - 0.38) | -0.03 | (-0.09 - 0.02) | -0.03 | (-0.06 - 0.01) | -0.03 | (-0.07 - 0.02) |
| Pandemic period (Mar-Dec 2020) | | | | | | | | |
| March 2020 | 0.23*** | (0.06 - 0.40) | -0.08*** | (-0.14 - -0.02) | -0.06*** | (-0.10 - -0.02) | -0.06** | (-0.10 - -0.01) |
| April 2020 | 0.13 | (-0.04 - 0.30) | -0.07** | (-0.13 - -0.01) | 0.07*** | (0.03 - 0.11) | 0.00 | (-0.05 - 0.05) |
| May 2020 | -0.13 | (-0.30 - 0.05) | -0.14*** | (-0.20 - -0.08) | 0.01 | (-0.02 - 0.05) | -0.03 | (-0.08 - 0.02) |
| June 2020 | -0.14 | (-0.32 - 0.03) | -0.12*** | (-0.18 - -0.05) | 0.02 | (-0.02 - 0.06) | 0.02 | (-0.03 - 0.07) |
| July 2020 | -0.01 | (-0.18 - 0.17) | -0.04 | (-0.10 - 0.02) | 0.08*** | (0.04 - 0.12) | 0.07*** | (0.02 - 0.12) |
| August 2020 | -0.02 | (-0.20 - 0.15) | -0.02 | (-0.09 - 0.04) | 0.07*** | (0.04 - 0.11) | 0.07*** | (0.02 - 0.12) |
| September 2020 | -0.14 | (-0.32 - 0.03) | -0.05 | (-0.11 - 0.01) | 0.04** | (0.00 - 0.08) | 0.04 | (-0.01 - 0.09) |
| October 2020 | -0.20** | (-0.38 - -0.03) | -0.10*** | (-0.16 - -0.04) | 0.02 | (-0.01 - 0.06) | 0.03 | (-0.02 - 0.08) |
| November 2020 | -0.21** | (-0.39 - -0.04) | -0.08*** | (-0.14 - -0.02) | 0.03 | (-0.01 - 0.07) | 0.00 | (-0.04 - 0.05) |
| December 2020 | -0.32*** | (-0.49 - -0.14) | -0.16*** | (-0.22 - -0.10) | 0.02 | (-0.02 - 0.05) | 0.05** | (0.01 - 0.10) |
| Pandemic x |  |  |  |  |  |  |  |  |
| Age < 15 |  |  |  |  |  |  |  |  |
| Age 15-24 |  |  |  |  | 0.04 | (-0.10 - 0.19) |  |  |
| Age 25-44 | 0.12 | (-0.06 - 0.31) | 0.20*** | (0.09 - 0.30) | 0.12*** | (0.07 - 0.16) | 0.07 | (-0.02 - 0.15) |
| Age 45-64 | 0.12* | (-0.01 - 0.25) | 0.10*** | (0.04 - 0.15) | 0.04*** | (0.01 - 0.07) | 0.06*** | (0.02 - 0.10) |
| Age 65-74 | 0.20*** | (0.06 - 0.35) | 0.03 | (-0.03 - 0.09) | 0.03* | (-0.01 - 0.06) | 0.05** | (0.01 - 0.09) |
| Age 75+ | *(omitted)* |  | *(omitted)* |  | *(omitted)* |  | *(omitted)* |  |
| Alpha | 0.030 |  | 0.003 |  | 0.001 |  | 0.001 |  |
| Constant | 0.90*** | (0.84 - 0.97) | 0.41*** | (0.32 - 0.50) | 0.63*** | (0.56 - 0.70) | 0.37*** | (0.30 - 0.44) |
| Observations | 1,584 |  | 1,848 |  | 2,112 |  | 2,640 |  |

**eTable 3 (Continued)**

|  | **Cancer** | | **Alzheimer's Disease** | | **Diabetes Mellitus** | | **External Causes** | |
| --- | --- | --- | --- | --- | --- | --- | --- | --- |
|  |  |  | Coef | 95% CI | Coef | 95% CI | Coef | 95% CI |
| Pre-pandemic portion of 2020 | | | | | | | | |
| January 2020 | 0.00 | (-0.02 - 0.01) | -0.11** | (-0.20 - -0.02) | -0.01 | (-0.06 - 0.04) | -0.04* | (-0.08 - 0.00) |
| February 2020 | 0.03*** | (0.01 - 0.04) | -0.02 | (-0.11 - 0.07) | 0.01 | (-0.04 - 0.07) | 0.01 | (-0.04 - 0.05) |
| Pandemic period (Mar-Dec 2020) | | | | | | | | |
| March 2020 | -0.01 | (-0.03 - 0.00) | 0.01 | (-0.08 - 0.10) | 0.07*** | (0.02 - 0.12) | -0.09*** | (-0.14 - -0.04) |
| April 2020 | -0.02** | (-0.04 - -0.00) | 0.21*** | (0.11 - 0.30) | 0.28*** | (0.22 - 0.33) | -0.11*** | (-0.16 - -0.06) |
| May 2020 | -0.04*** | (-0.06 - -0.02) | 0.08* | (-0.01 - 0.18) | 0.19*** | (0.13 - 0.24) | -0.04 | (-0.09 - 0.01) |
| June 2020 | -0.03*** | (-0.04 - -0.01) | 0.07 | (-0.03 - 0.16) | 0.14*** | (0.09 - 0.20) | 0.00 | (-0.05 - 0.05) |
| July 2020 | -0.01 | (-0.03 - 0.01) | 0.14*** | (0.05 - 0.23) | 0.25*** | (0.19 - 0.30) | 0.00 | (-0.05 - 0.05) |
| August 2020 | 0.00 | (-0.02 - 0.02) | 0.17*** | (0.08 - 0.27) | 0.23*** | (0.18 - 0.28) | 0.01 | (-0.04 - 0.06) |
| September 2020 | -0.01 | (-0.02 - 0.01) | 0.11** | (0.02 - 0.21) | 0.21*** | (0.15 - 0.26) | 0.00 | (-0.05 - 0.05) |
| October 2020 | -0.02** | (-0.04 - -0.00) | 0.04 | (-0.05 - 0.14) | 0.17*** | (0.12 - 0.22) | -0.01 | (-0.06 - 0.05) |
| November 2020 | -0.03*** | (-0.05 - -0.01) | 0.07 | (-0.02 - 0.17) | 0.23*** | (0.17 - 0.28) | -0.02 | (-0.07 - 0.03) |
| December 2020 | -0.01 | (-0.03 - 0.01) | 0.10** | (0.01 - 0.20) | 0.23*** | (0.18 - 0.29) | -0.03 | (-0.09 - 0.02) |
| Pandemic x |  |  |  |  |  |  |  |  |
| Age < 15 | -0.02 | (-0.14 - 0.10) |  |  |  |  | 0.03 | (-0.03 - 0.10) |
| Age 15-24 | 0.01 | (-0.12 - 0.13) |  |  |  |  | 0.15*** | (0.10 - 0.21) |
| Age 25-44 | -0.03* | (-0.07 - 0.00) |  |  | 0.04 | (-0.03 - 0.12) | 0.17*** | (0.12 - 0.21) |
| Age 45-64 | 0.00 | (-0.01 - 0.02) | -0.08 | (-0.20 - 0.04) | -0.04* | (-0.09 - 0.00) | 0.09*** | (0.05 - 0.14) |
| Age 65-74 | 0.03*** | (0.01 - 0.05) | 0.06 | (-0.02 - 0.13) | -0.02 | (-0.06 - 0.03) | -0.01 | (-0.07 - 0.04) |
| Age 75+ | *(omitted)* |  | *(omitted)* |  | *(omitted)* |  | *(omitted)* |  |
| Alpha | 0.000 |  | 0.006 |  | 0.002 |  | 0.003 |  |
| Constant | 0.76*** | (0.67 - 0.86) | 0.74*** | (0.66 - 0.81) | 0.24*** | (0.16 - 0.31) | 2.53*** | (2.48 - 2.59) |
| Observations | 2,640 |  | 1,056 |  | 1,848 |  | 2,640 |  |

* p<0.05, ** p<0.01, *** p<0.001

Note: All models also controlled for the main effects of age (<5,5-14,25-34,…75-84,85+), month, and calendar year (linear specification for influenza/pneumonia, cubic specification for other respiratory diseases and cancer; quadratic specification for the remaining outcomes). In addition, the models included interactions between the age groups and the time trend (i.e., linear, quadratic or cubic). The model for external causes also included interactions between month and broad age groups (i.e., <5, 5-34, 35+).

eTable 4. RMSE^a^ for alternative specifications of the pre-pandemic time trend

|  | **Model 1**  (1999-,  linear  time trend) | **Model 2**  (1999-,  quadratic  time trend) | **Model 3**  (1999-,  cubic  time trend) | **Model 4**  (2009-,  linear  time trend) | **Model 5**  (2014-,  linear  time trend) |
| --- | --- | --- | --- | --- | --- |
| **Overall RMSE** |  |  |  |  |  |
| All causes | 768 | 703 | 695 | 706 | 736 |
| Influenza/pneumonia | 105 | 104 | 104 | 103 | 108 |
| Other respiratory disease | 85 | 82 | 81 | 74 | 69 |
| Heart disease | 291 | 209 | 197 | 172 | 160 |
| Cerebrovascular disease | 112 | 67 | 61 | 59 | 49 |
| Cancer | 74 | 61 | 58 | 75 | 64 |
| Alzheimer’s disease | 112 | 113 | 107 | 118 | 120 |
| Diabetes | 34 | 30 | 27 | 25 | 24 |
| External causes | 76 | 66 | 60 | 65 | 67 |
| **RMSE for 2019** |  |  |  |  |  |
| All causes | 596 | 309 | 402 | 403 | 345 |
| Influenza/pneumonia | 53 | 55 | 57 | 71 | 83 |
| Other respiratory disease | 99 | 84 | 66 | 77 | 66 |
| Heart disease | 385 | 107 | 148 | 133 | 100 |
| Cerebrovascular disease | 158 | 43 | 71 | 60 | 37 |
| Cancer | 98 | 66 | 57 | 92 | 65 |
| Alzheimer’s disease | 134 | 76 | 198 | 105 | 112 |
| Diabetes | 48 | 26 | 27 | 24 | 24 |
| External causes | 95 | 57 | 92 | 72 | 74 |

Note: Models 1-3 were fit to the data for 1999-2020; the only difference between them is that the specification of the time trend was linear in Model 1, quadratic in Model 3, and cubic in Model 3. The remaining parameters were specified as shown in Eq. (1) except that the interactions between the month dummies and age group (<5, 5-34, 35) were included only for all causes and external causes. Models 4 and 5 were fit to a shorter data series (2009-2020 for Model 4 and 2014-2020 for Model 5). Both of those models used a specification similar to Model 1 (i.e., linear time trend), except that the time trend was interacted with broader age groups (<15, 15-24, 25-44, 45-64, 65-74, 75+) rather than the most detailed age dummies (<5, 5-14,….75-84, 85+). This simplification was necessary because the shorter time series did not provide sufficient variation to identify all the parameters in a more complicated specification. For some outcomes, the model would not converge if the linear time trend was interacted with all age groups or if the time trend was specified as quadratic. The results presented in Tables 1 and 2 come from Model 2 for all outcomes except influenza/pneumonia (which is based on Model 1)

and two groups of causes that are based on Model 3 (i.e., other respiratory diseases and cancer).

^a^ The root mean squared error (RMSE)—which represents a measure of predictive accuracy—is computed as: $\sqrt{\frac{\sum_{i=1}^{N} \left( D-\hat{D} \right)^{2}}{N}}$, where $D$ represents the observed number of deaths; $\hat{D}$ denotes the predicted number of deaths; and $N$ represents the number of observations. Higher values indicate more error (i.e., larger residuals). Values are scale-dependent (i.e., they depend on the number of deaths from that cause and thus, will be highest for all-cause deaths) and cannot be compared across outcomes.

eFigure 1. Excess Mortality from External Causes by Age Group During March-December 2020, US, Females: a) Rate Ratios; B) Number of Excess Deaths

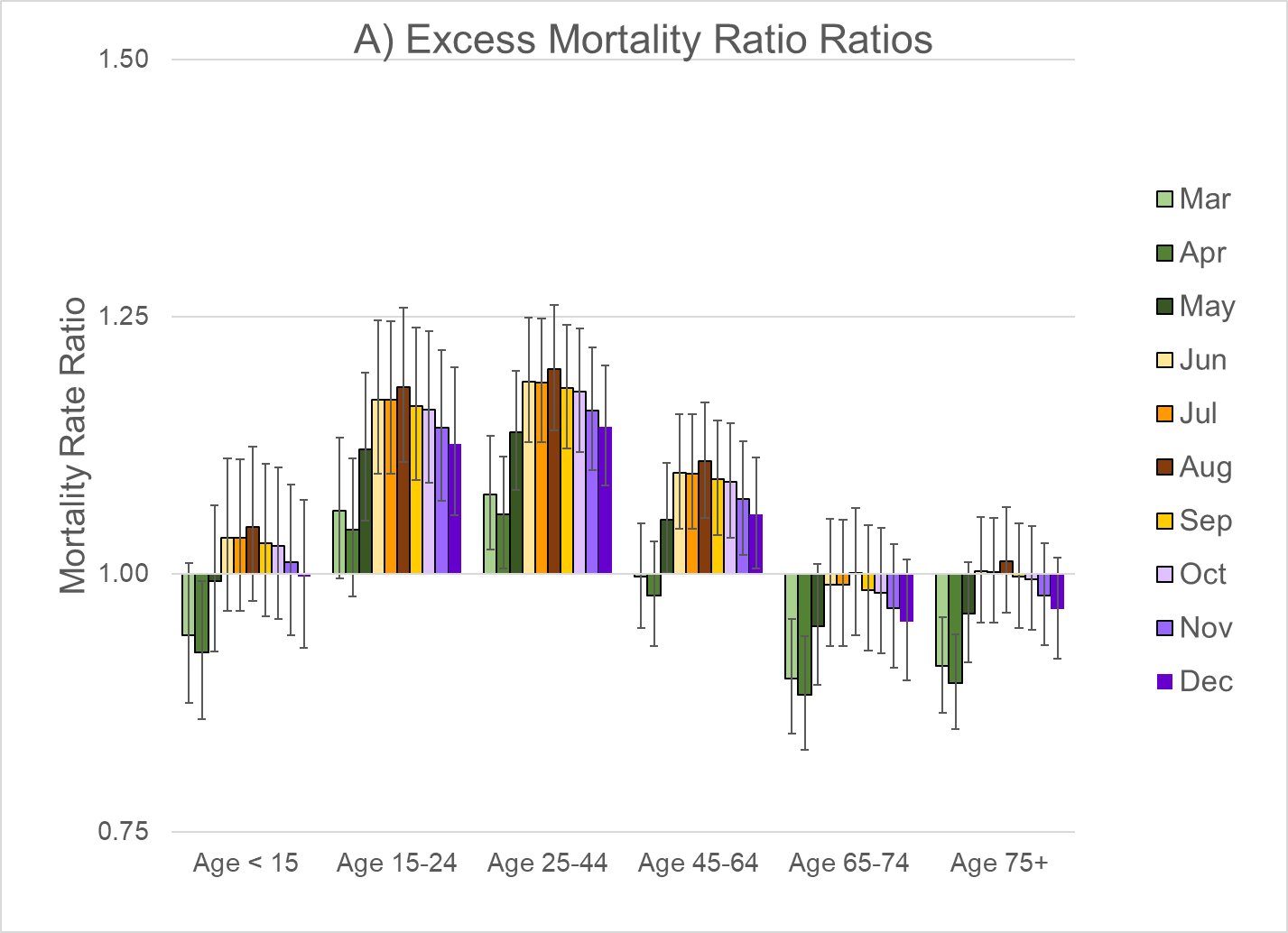

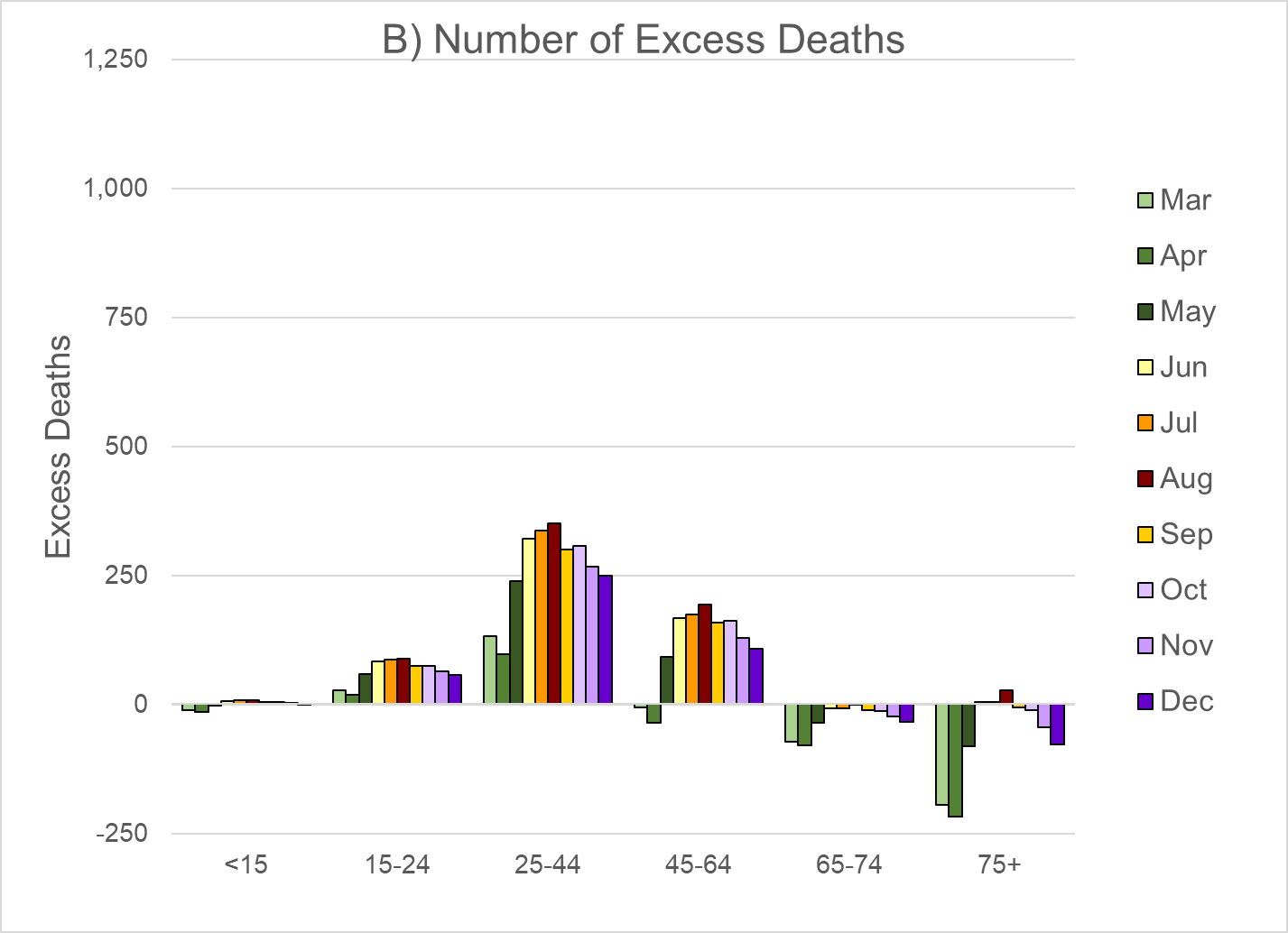

Note: These estimates are based on the regression models shown in eTable 3. The bars on the graph of excess mortality rate ratios represent the 95% confidence interval.

eFigure 2. Excess Mortality Rate Ratios for Selected Causes of Death by Age Group During March-December 2020, US, Males

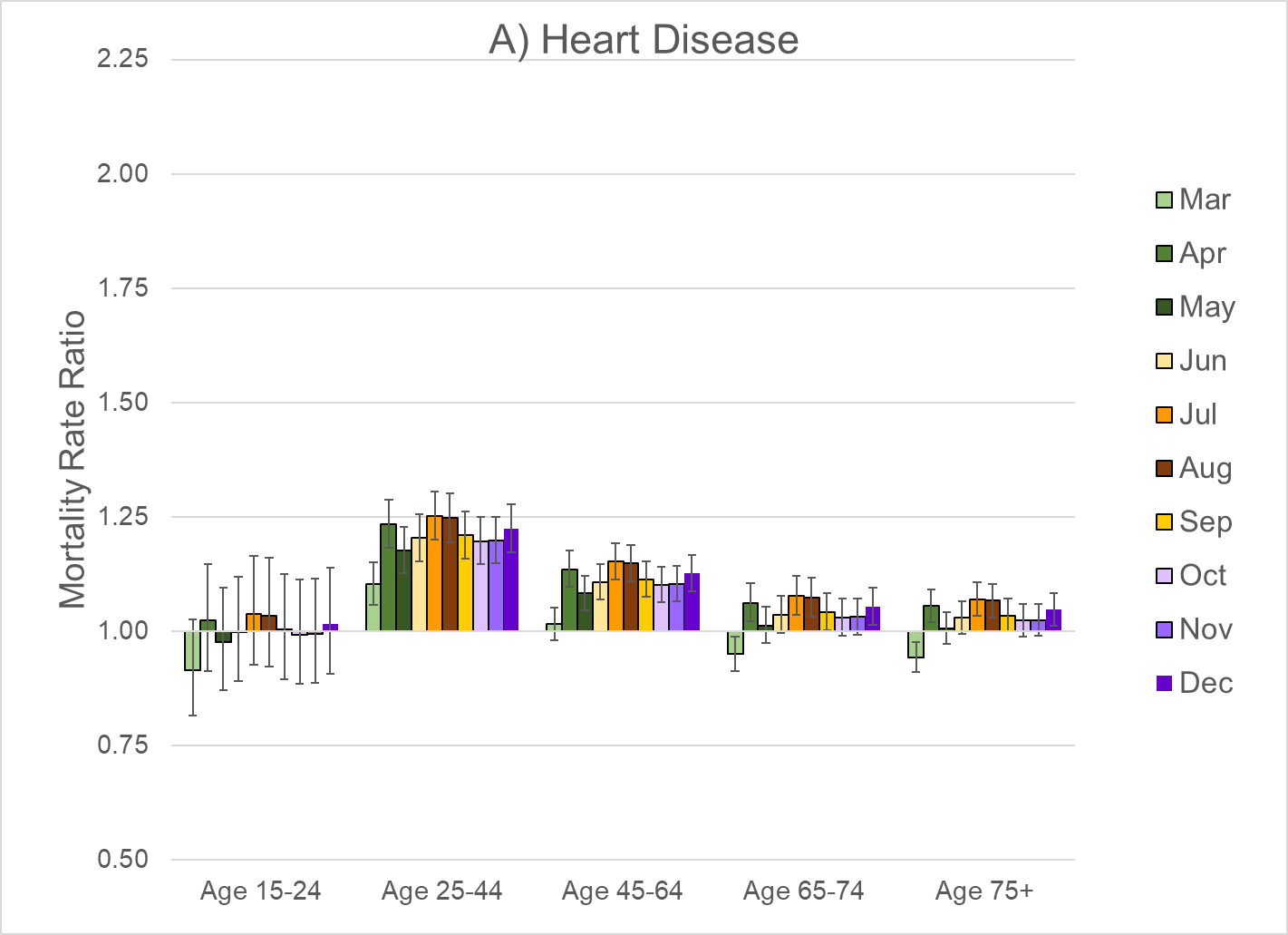

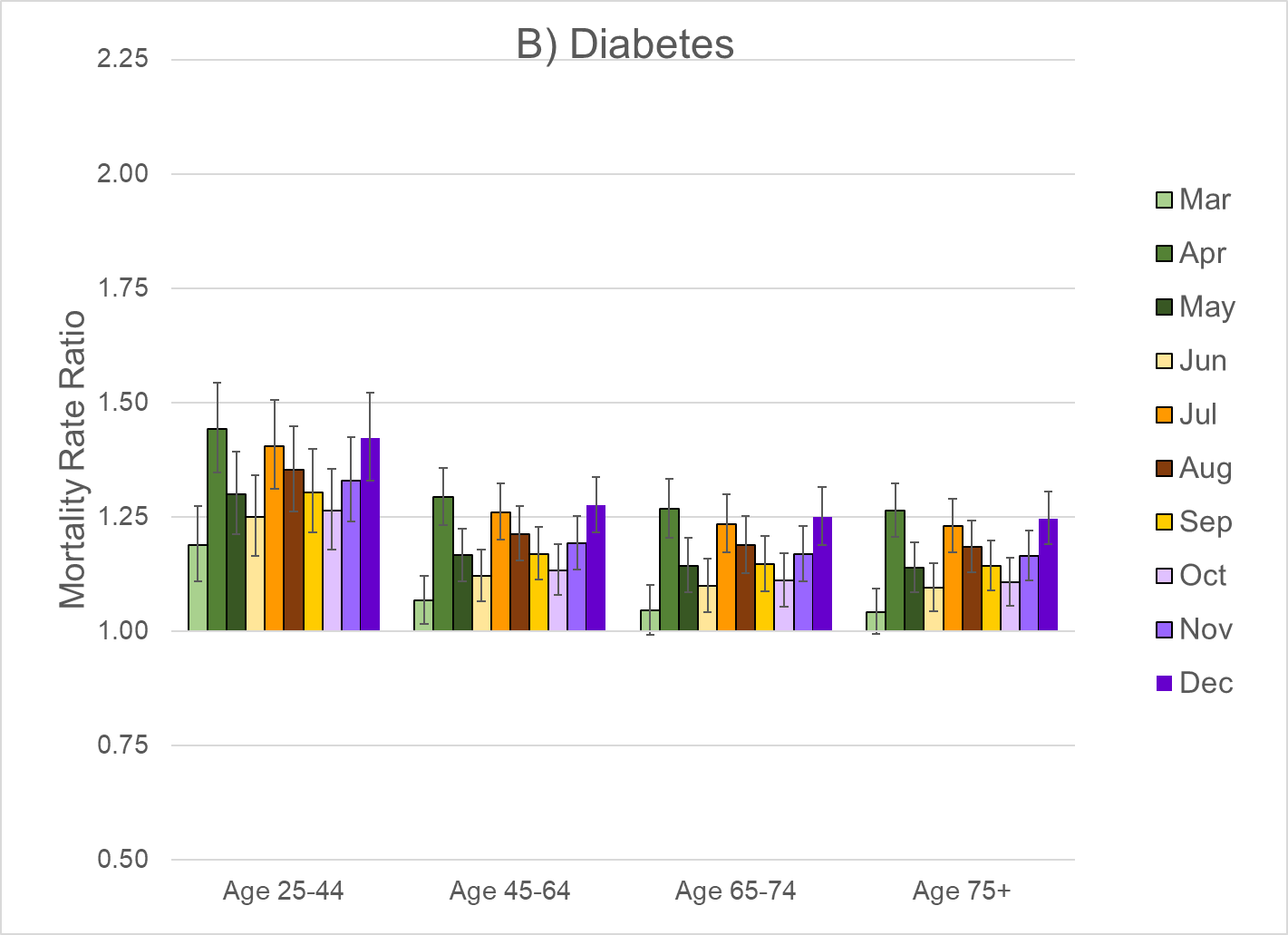

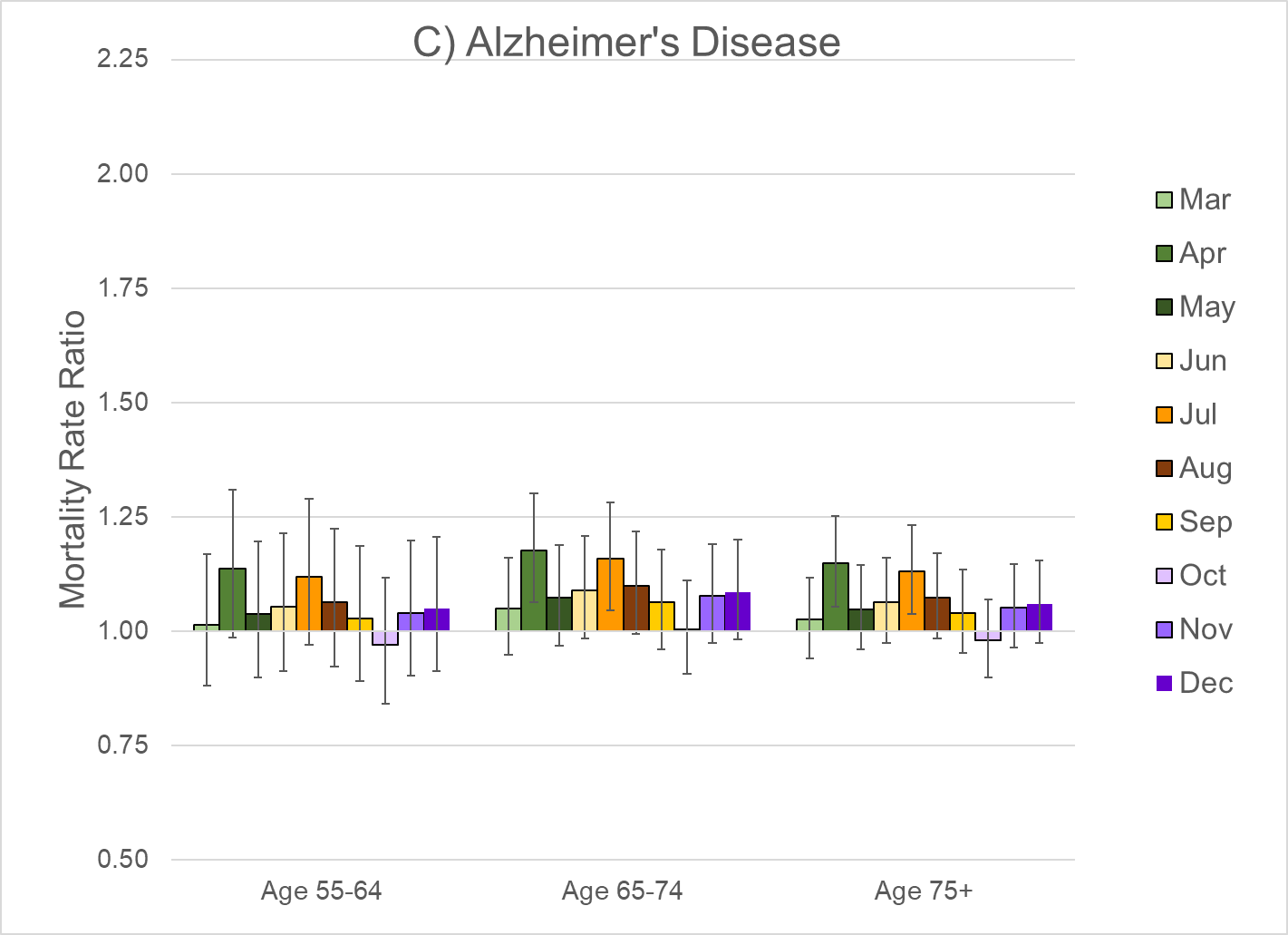

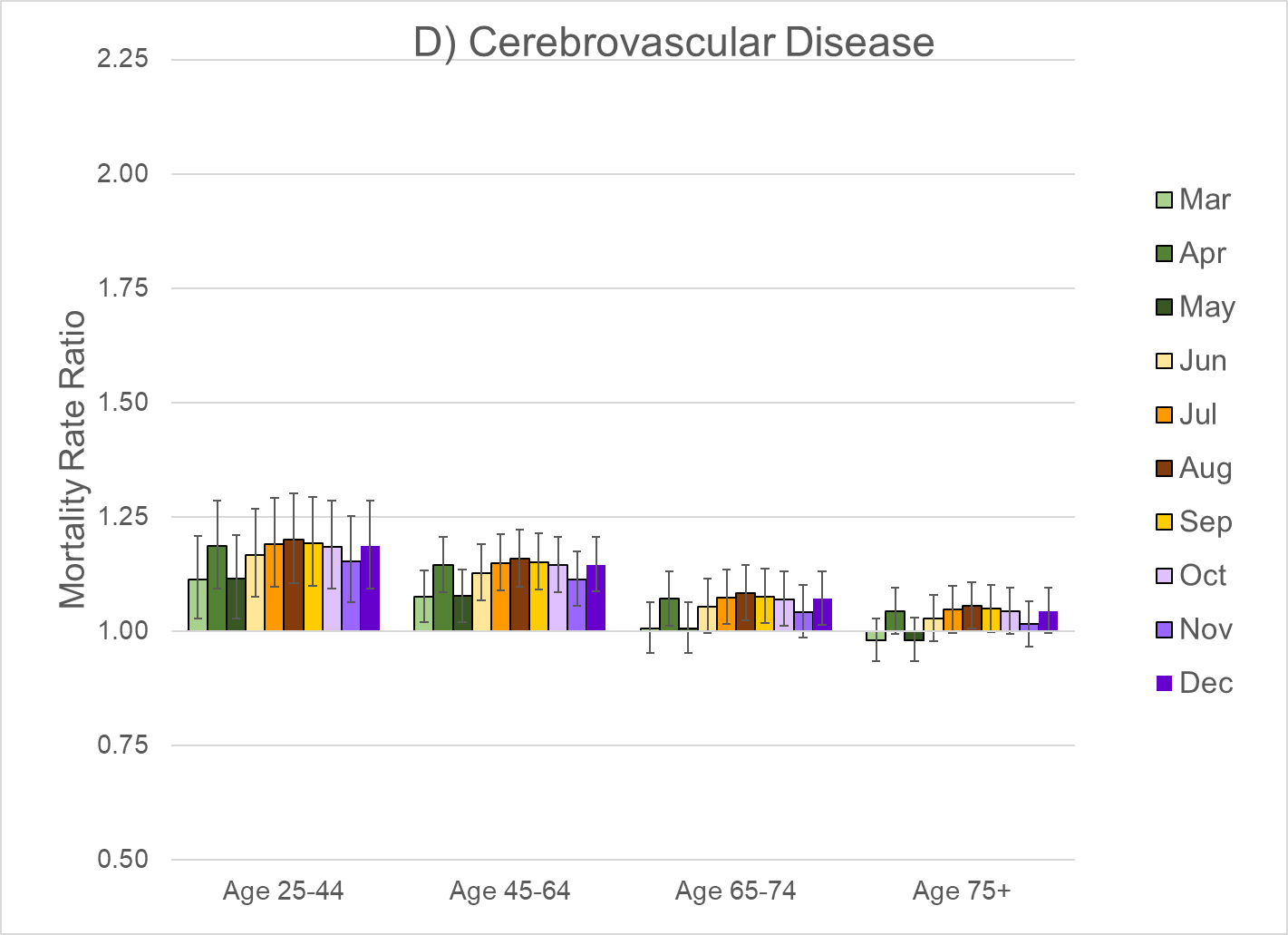

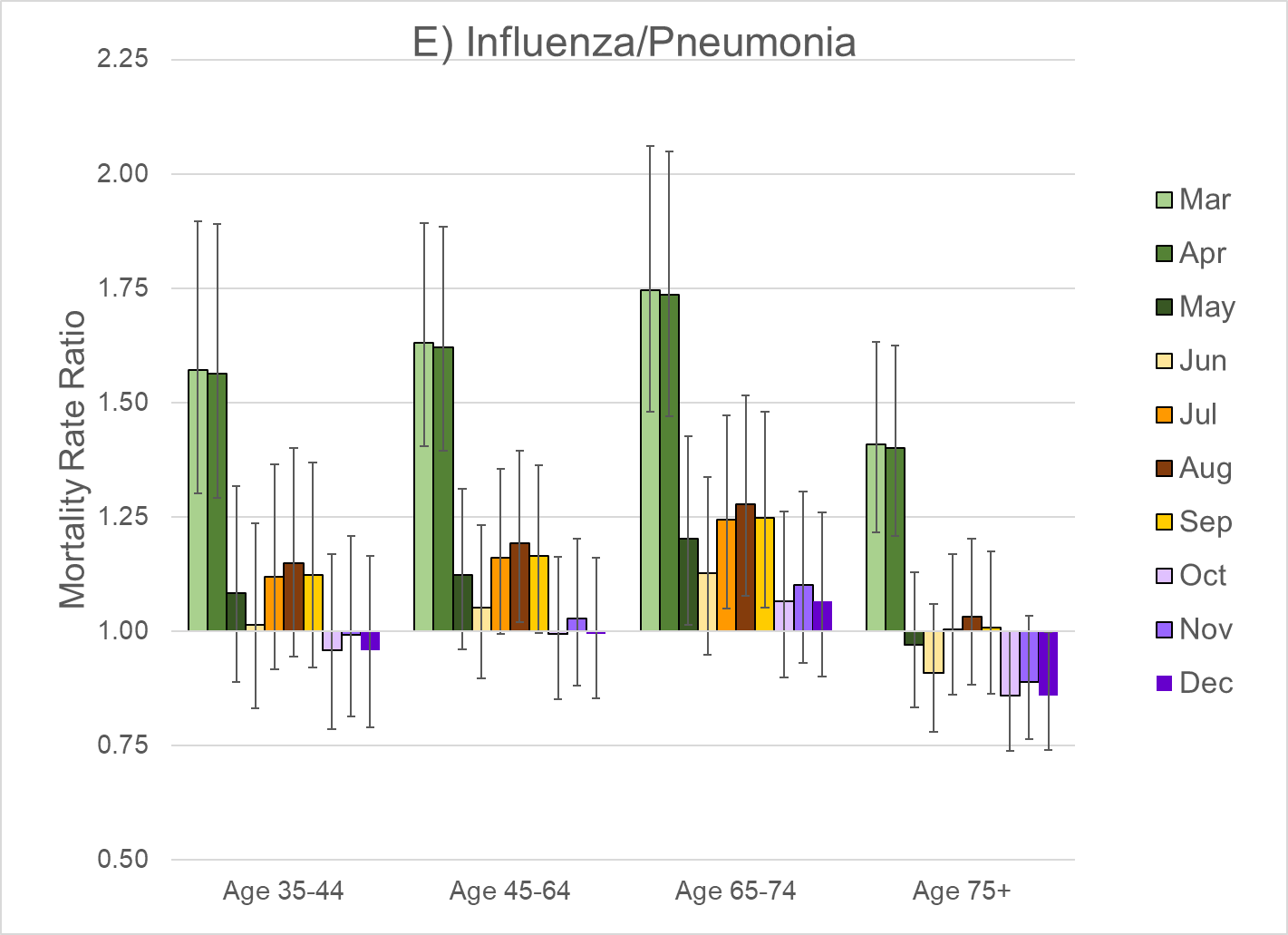

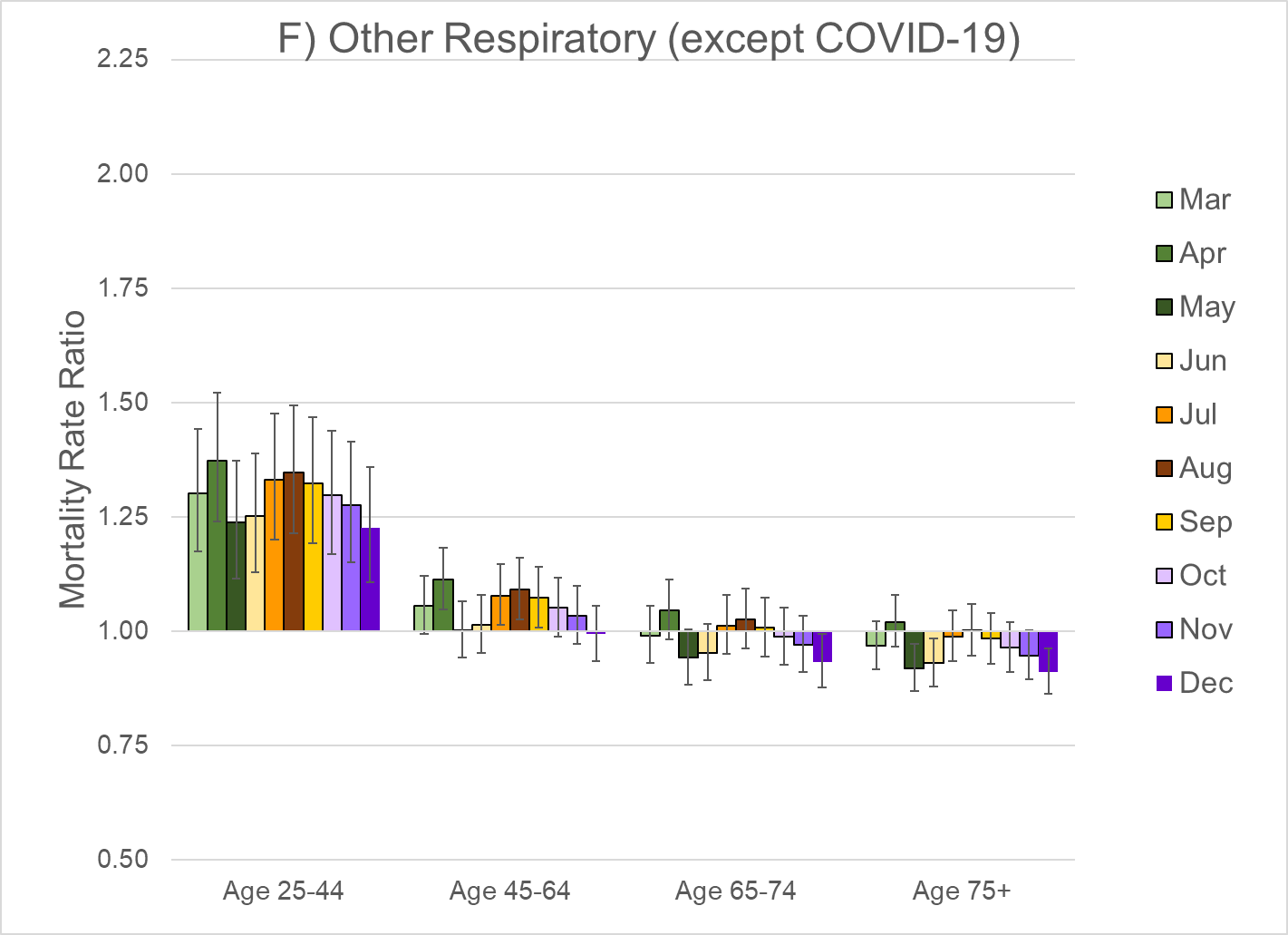

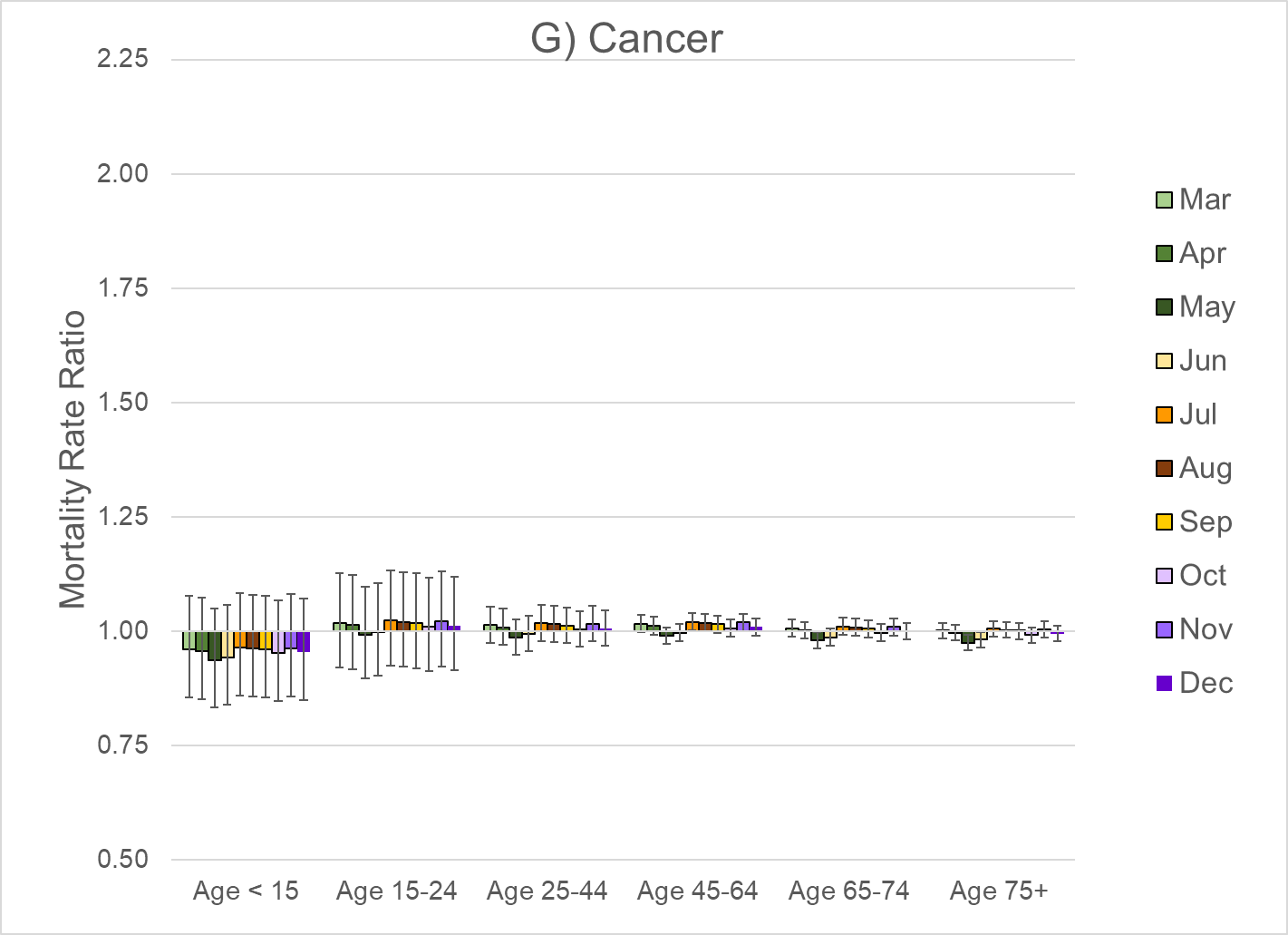

Note: These rate ratios are based on the regression models shown in eTable 2. The bars represent the 95% confidence interval.

eFigure 3. Excess Mortality Rate Ratios for Selected Causes of Death by Age Group During March-December 2020, US, Females

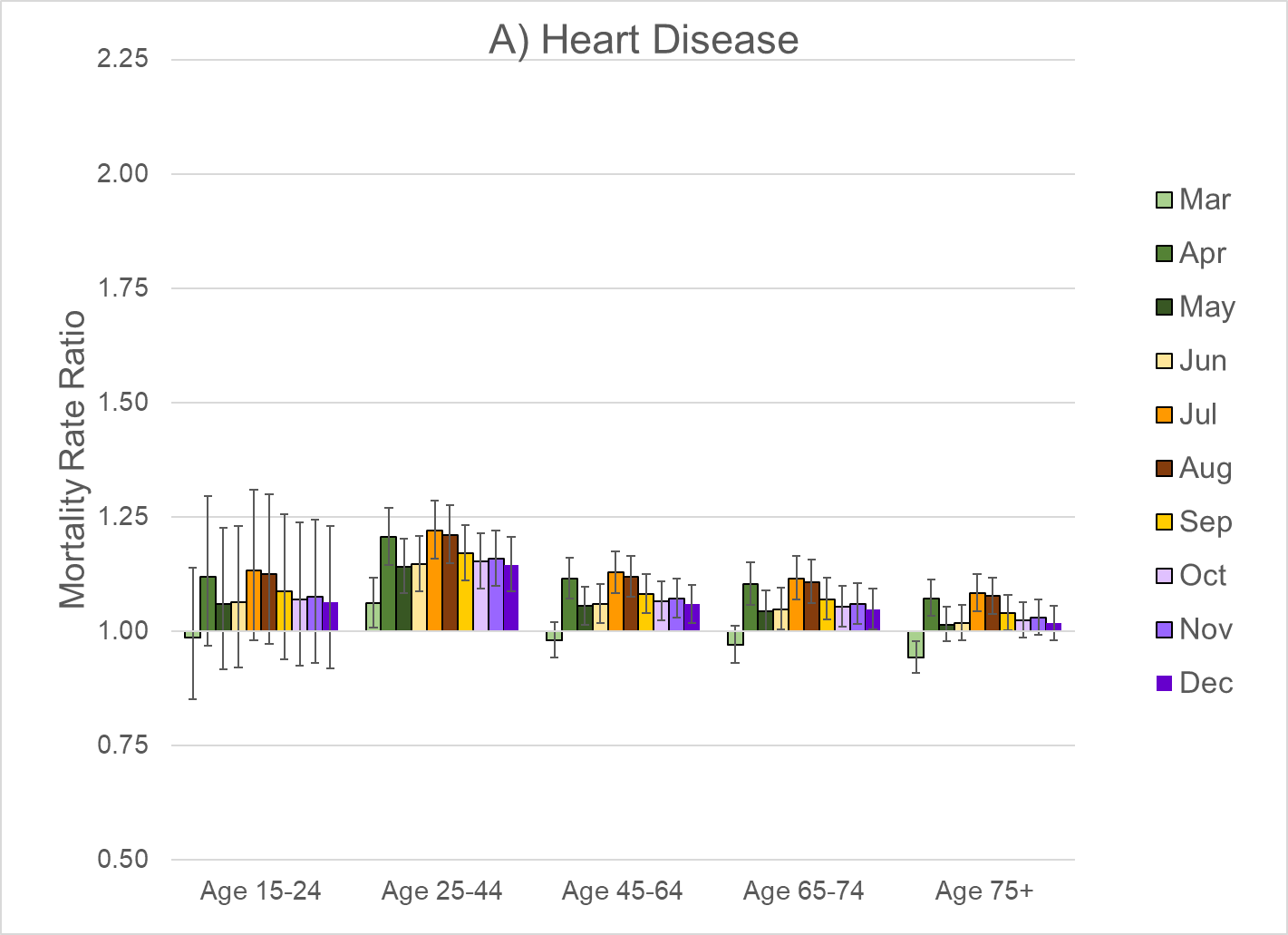

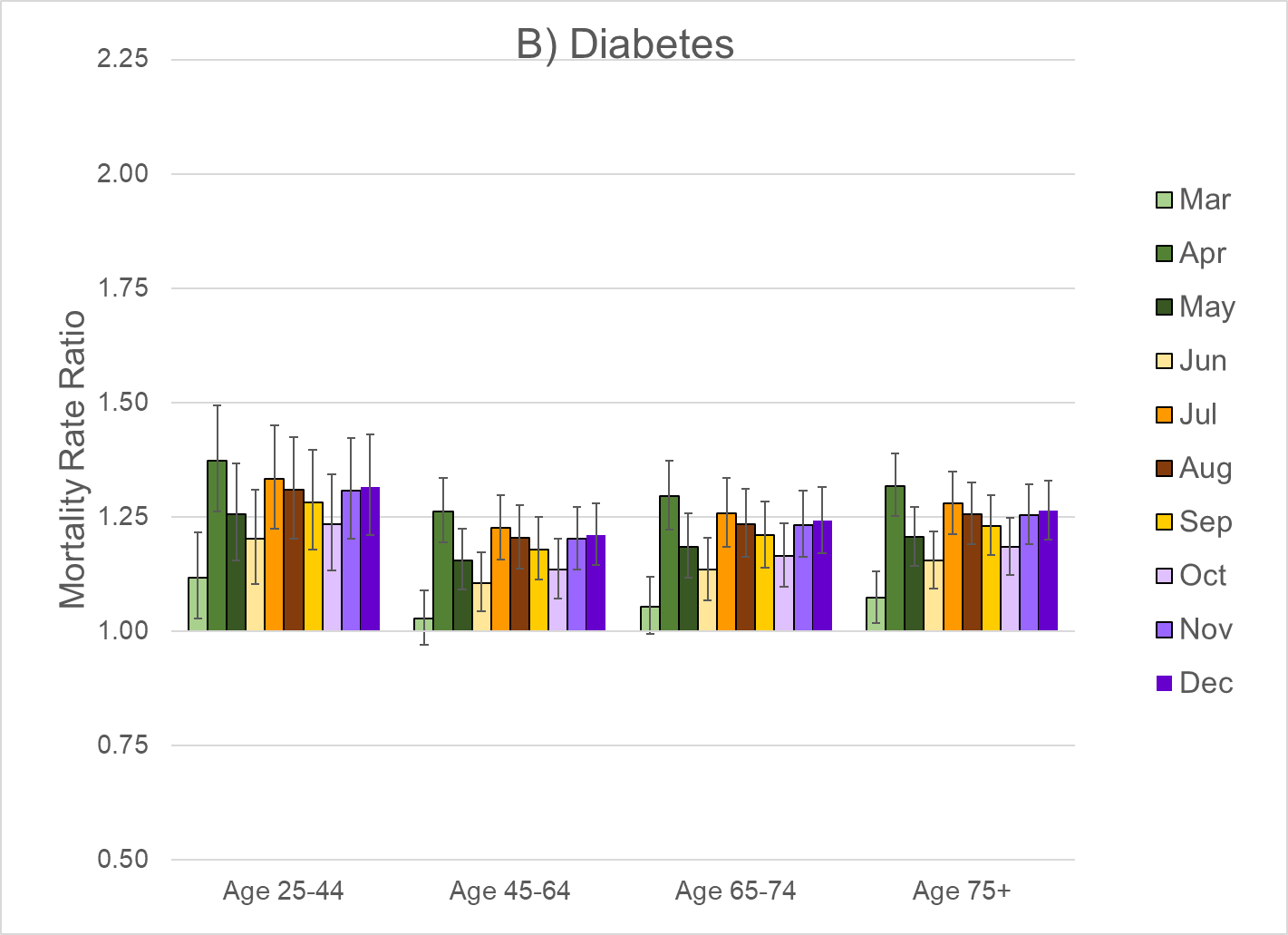

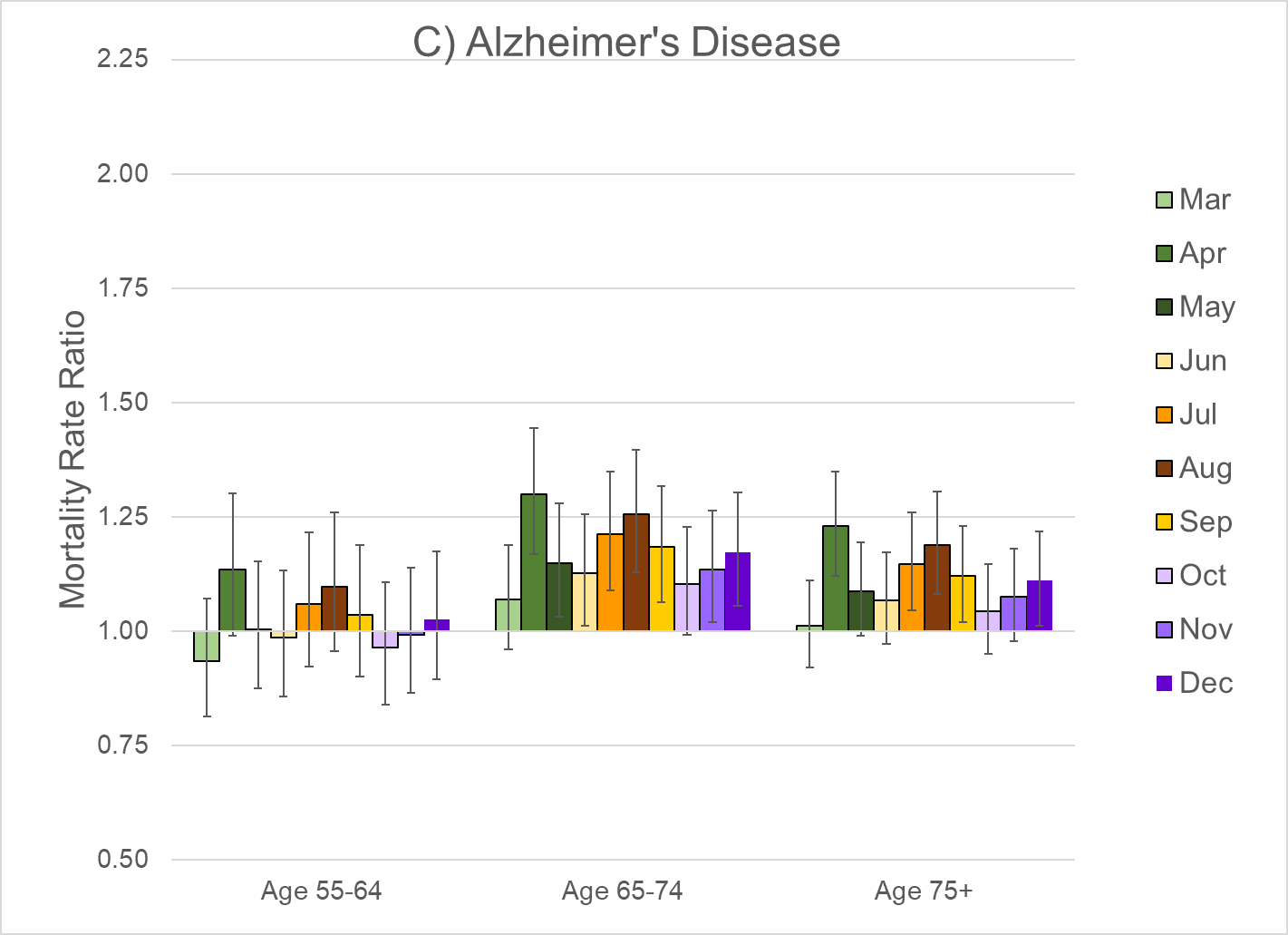

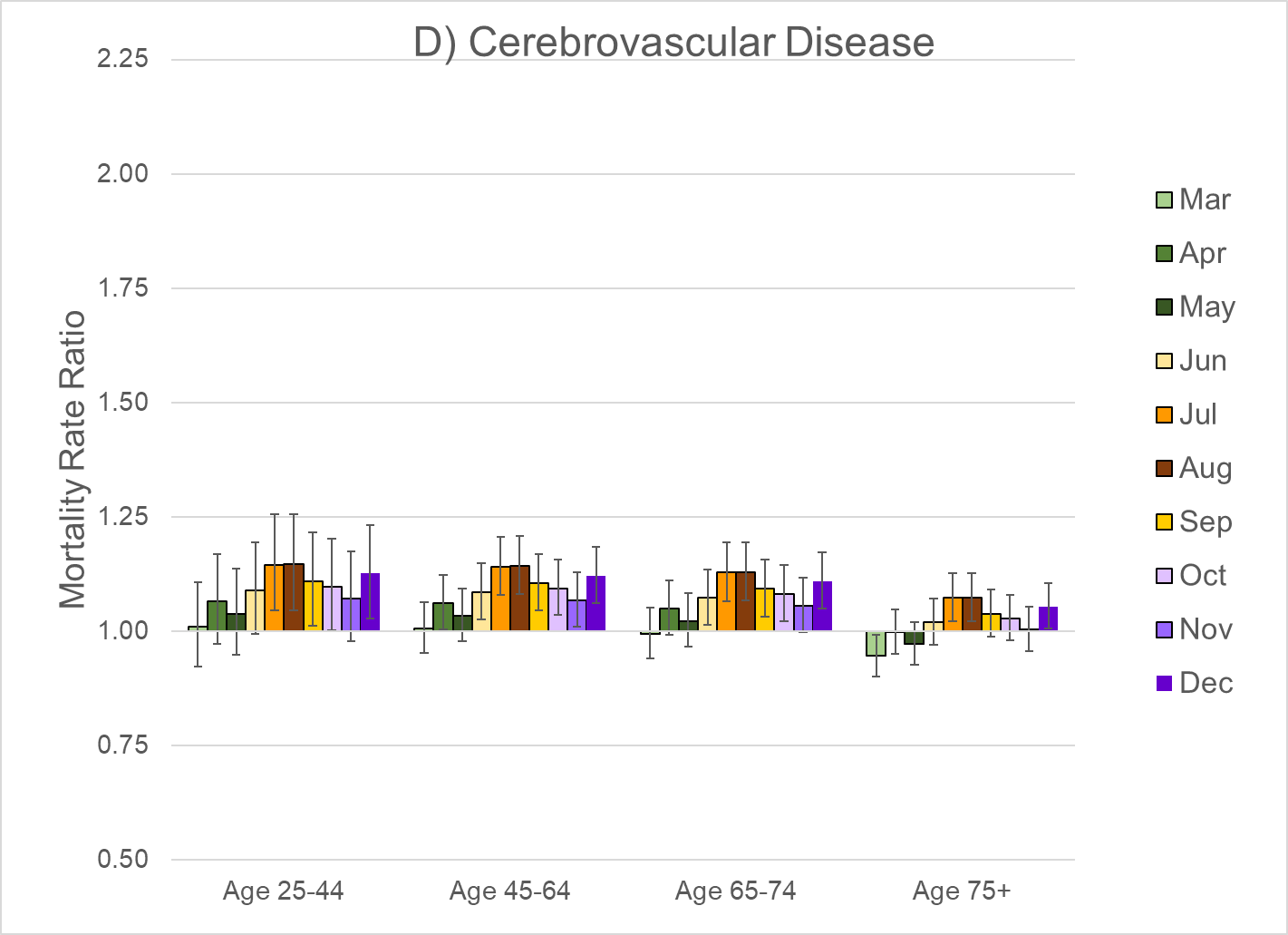

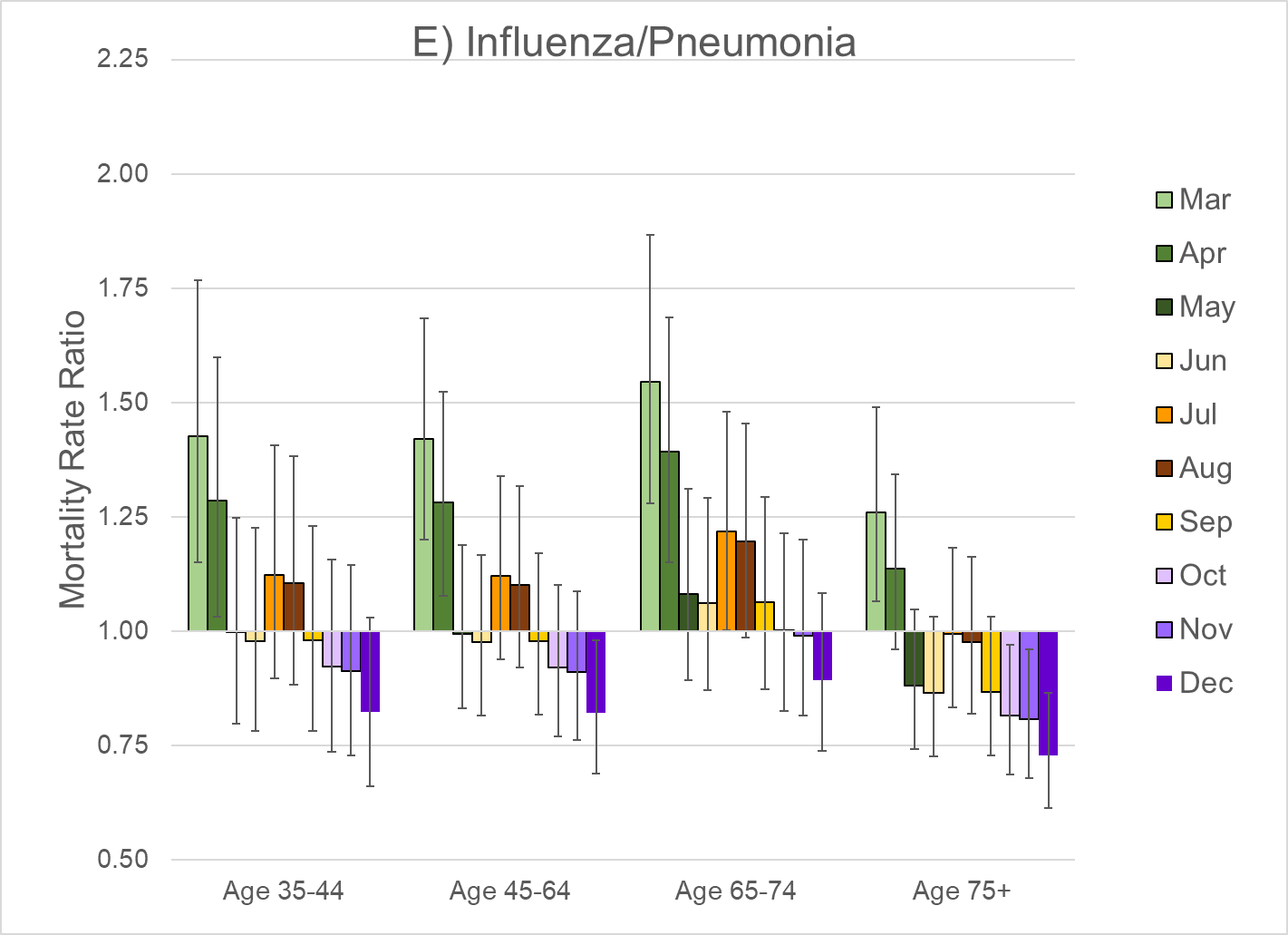

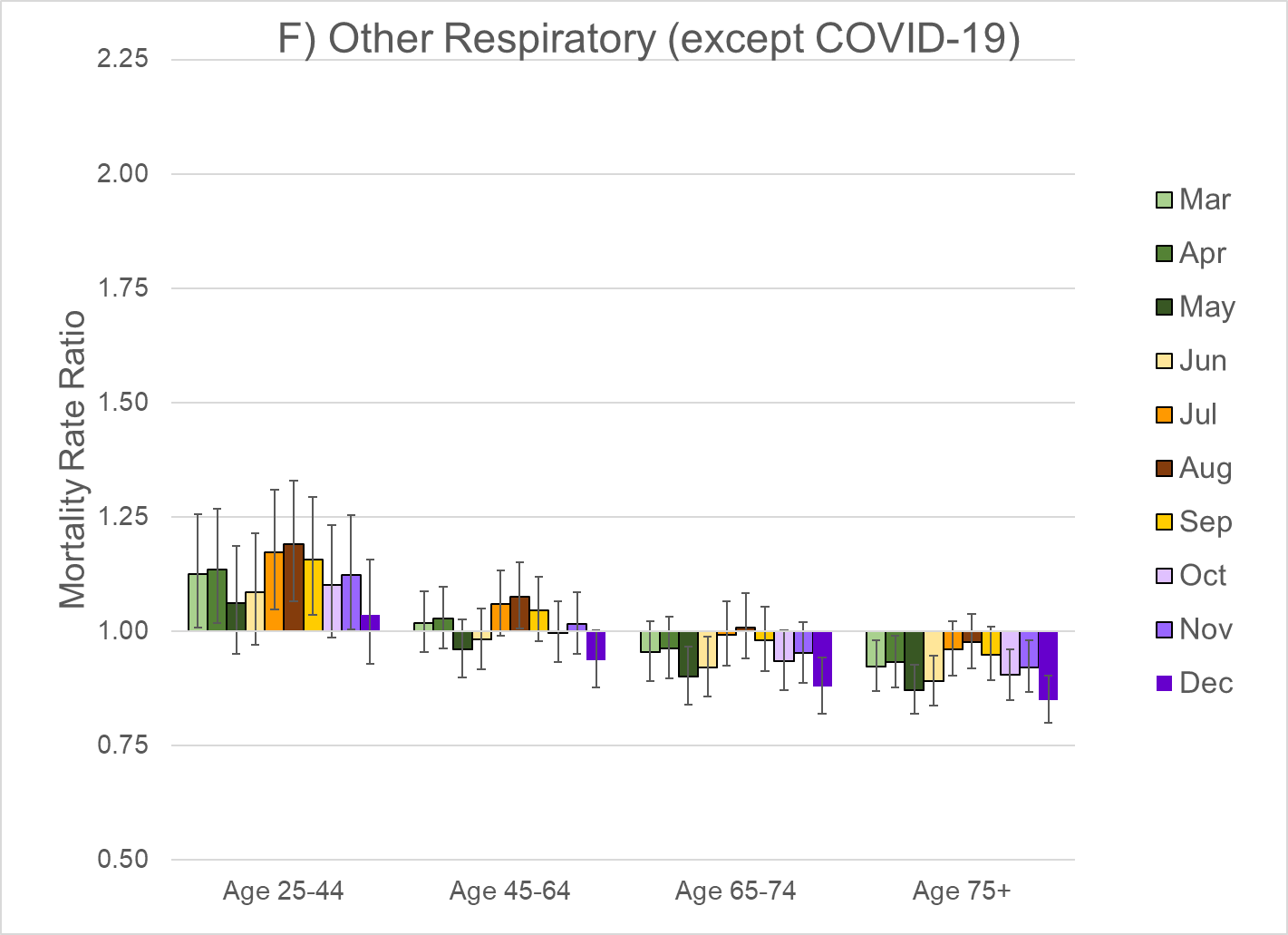

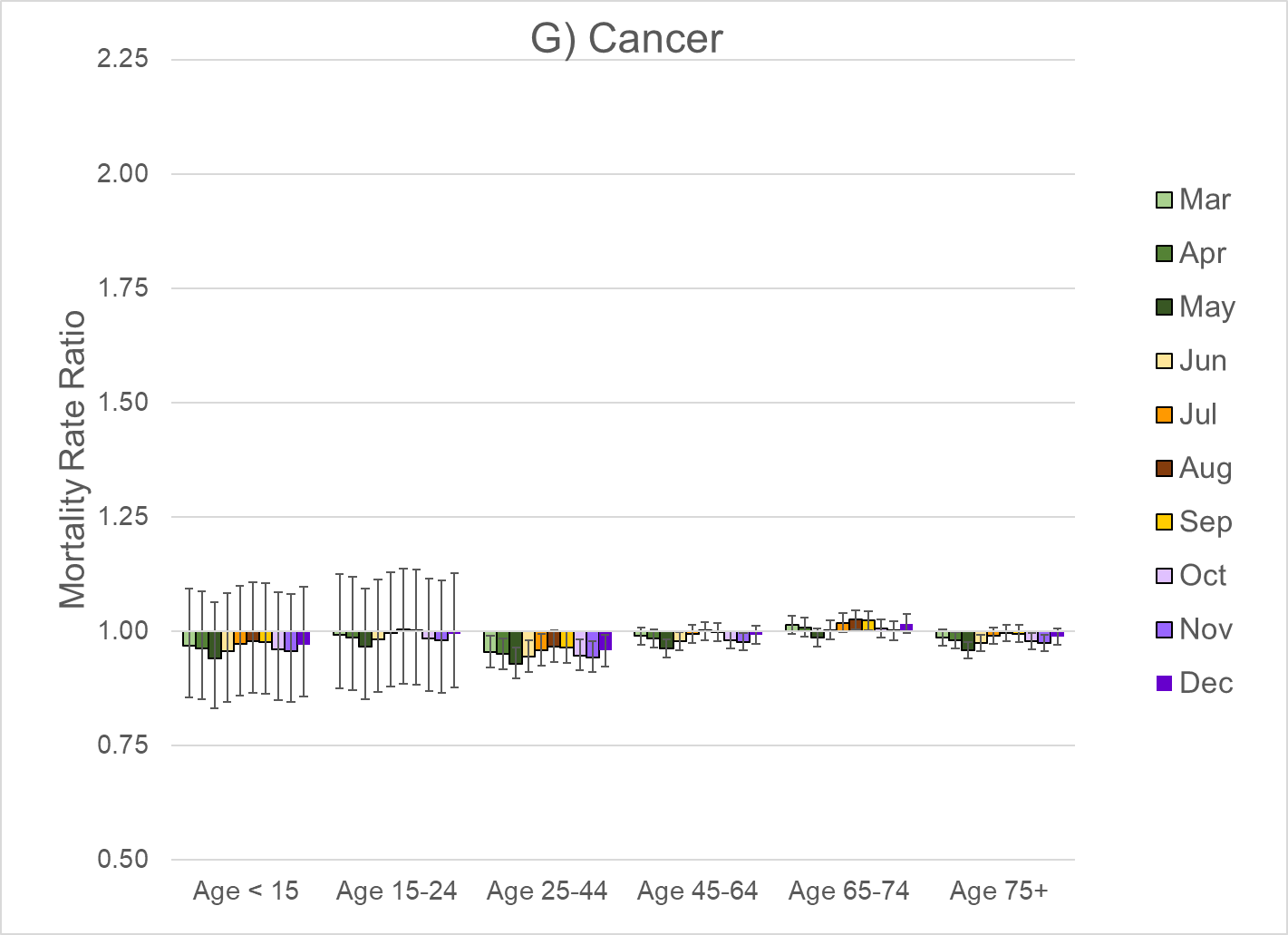

Note: These rate ratios are based on the regression models shown in eTable 3. The bars represent the 95% confidence interval.

eFigure 4. Estimated number of excess all-cause deaths during March-December 2020 for alternative specifications of the pre-pandemic time trend by sex

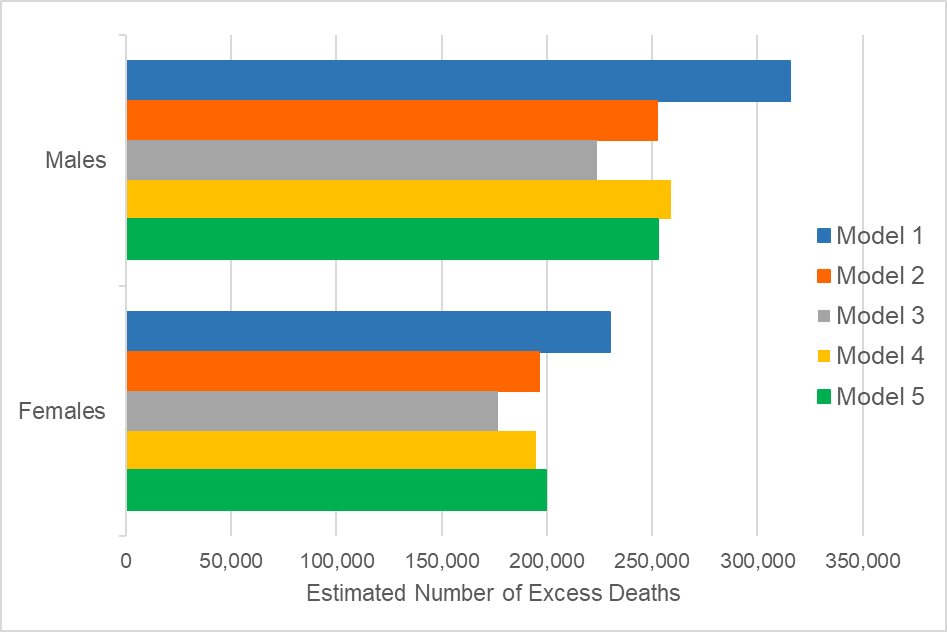

Note: The results from Model 2 match the results presented in Tables 1 and 2. Models 1-3 were fit to the data for 1999-2020, but the specification of the time trend was linear in Model 1, quadratic in Model 3, and cubic in Model 3. The remaining parameters were specified as shown in Eq. (1). Models 4 and 5 were fit to a shorter data series (2009-2020 for Model 4 and 2014-2020 for Model 5). Both of these models used a specification similar to Model 1 (i.e., linear time trend), except that the time trend was interacted with broader age groups (<15, 15-24, 25-44, 45-64, 65-74, 75+) rather than the most detailed age dummies (<5, 5-14,….75-84, 85+).

eFigure 5. Estimated number of excess deaths from specific causes during March‑December 2020 for alternative specifications of the pre-pandemic time trend: A) Males and B) Females

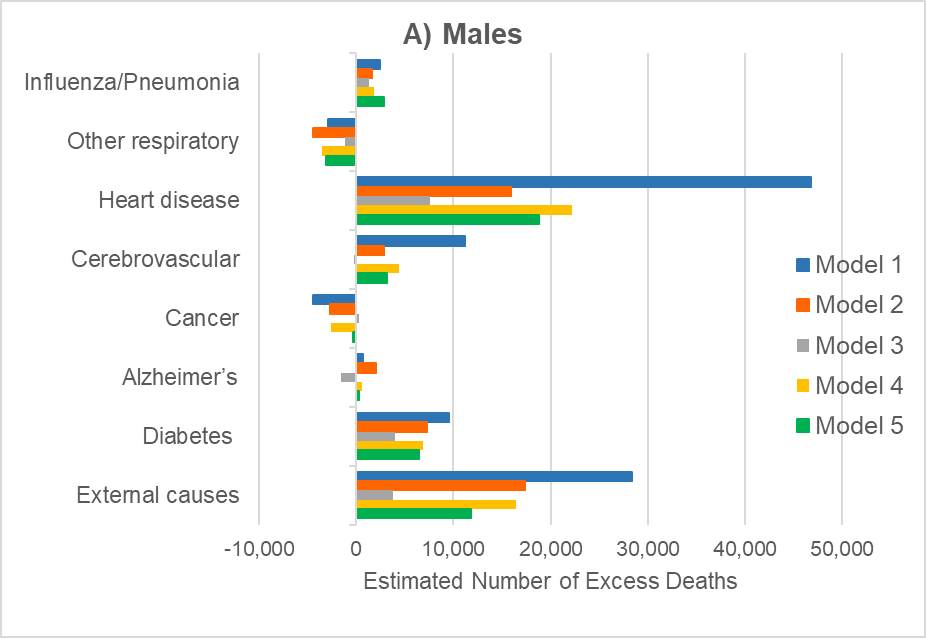

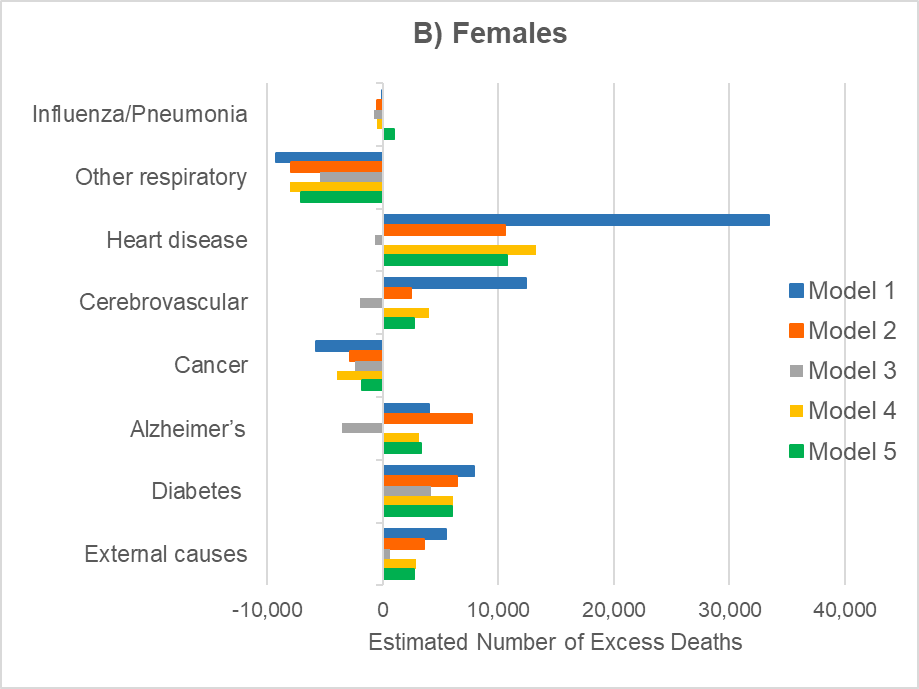

Note: Most of the cause-specific results presented in Tables 1 and 2 match Model 2. The exceptions are influenza/pneumonia (which comes from Model 1) and two groups of causes that are based on Model 3 (i.e., other respiratory diseases and cancer). Models 1-3 were fit to the data for 1999-2020, but the specification of the time trend was linear in Model 1, quadratic in Model 3, and cubic in Model 3. The remaining parameters were specified as shown in Eq. (1) except that the interactions between the month dummies and age group (<5, 5-34, 35) were included only for external causes. Models 4 and 5 were fit to a shorter data series (2009-2020 for Model 4 and 2014-2020 for Model 5). Both of these models used a specification similar to Model 1 (i.e., linear time trend), except that the time trend was interacted with broader age groups (<15, 15-24, 25-44, 45-64, 65-74, 75+) rather than the most detailed age dummies (<5, 5-14,….75-84, 85+).

eFigure 6. Observed versus expected all-cause deaths among men ages 65-74 during 2019-2021

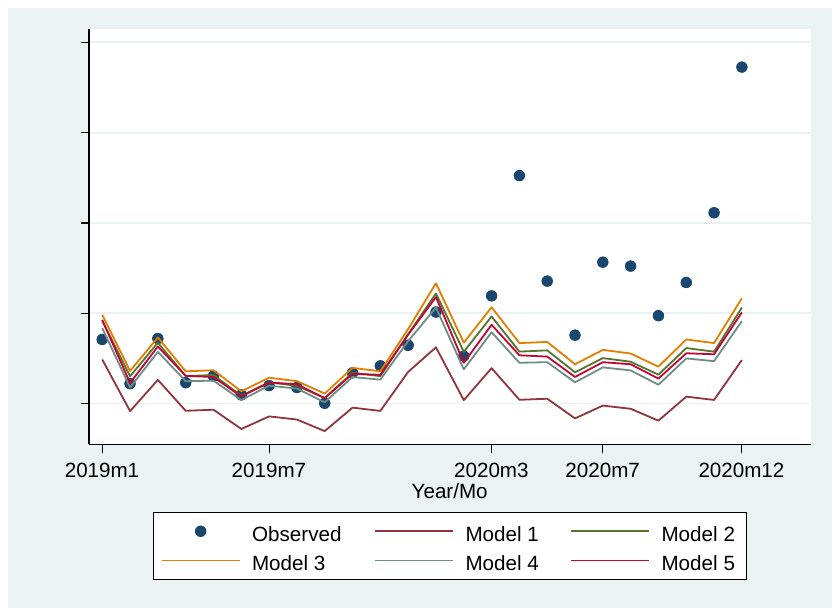

eFigure 7. Observed versus expected heart disease deaths among women aged 85 and older during 2019-2021

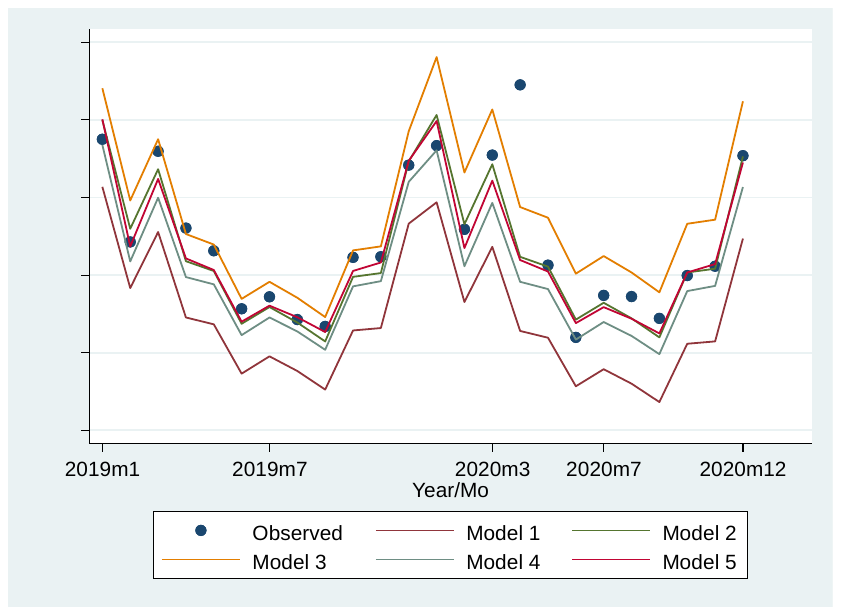

eFigure 8. Observed versus expected heart disease deaths during April among women aged 85 and older during 1999-2020

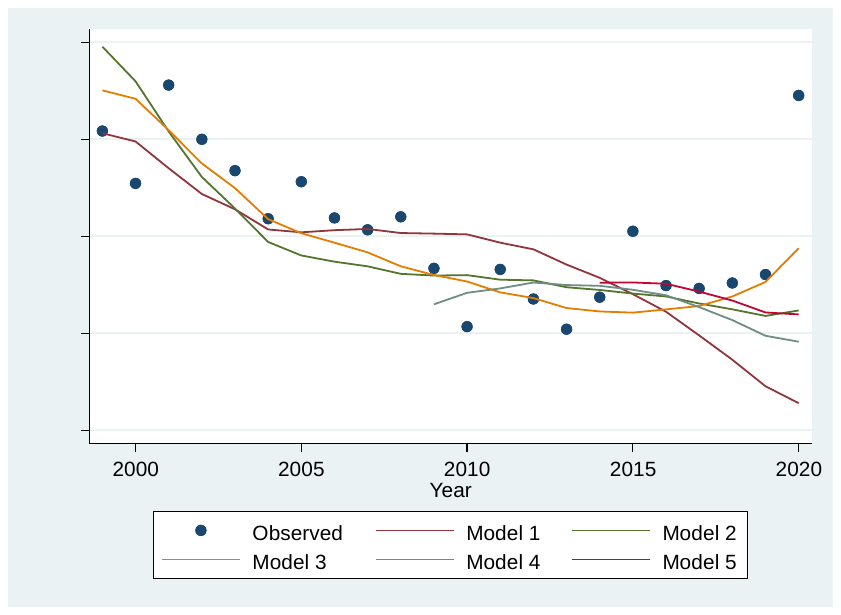

eFigure 9. Observed versus expected deaths from external causes among men aged 25‑34 during 2019-2021

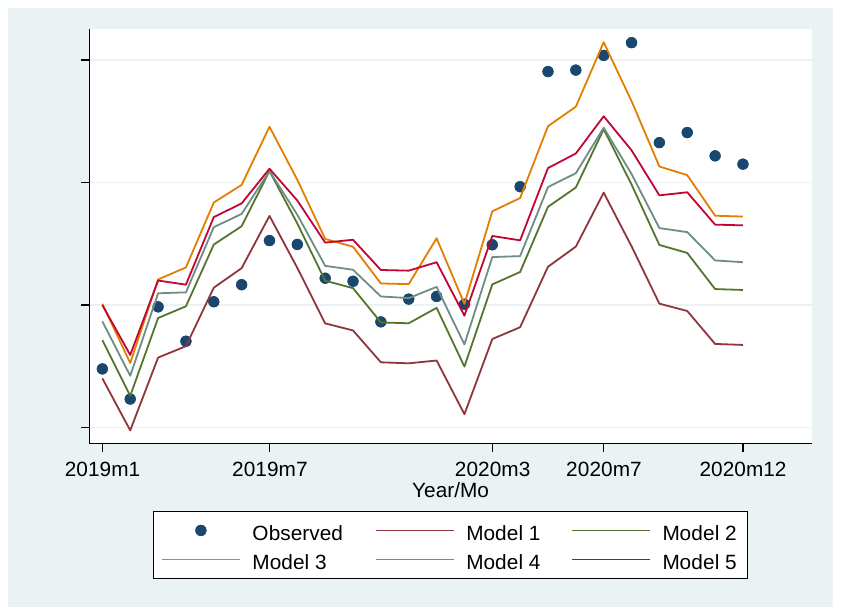

eFigure 10. Observed versus expected deaths from external causes during August among men aged 25-34 during 1999-2020

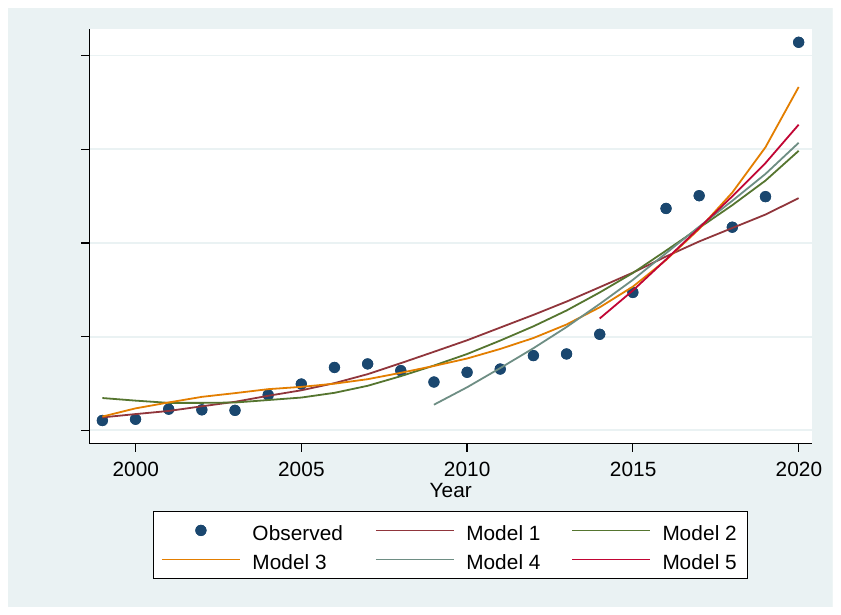

eFigure 11. Observed versus expected deaths from Alzheimer’s disease among women aged 85 and older during 2019-2021

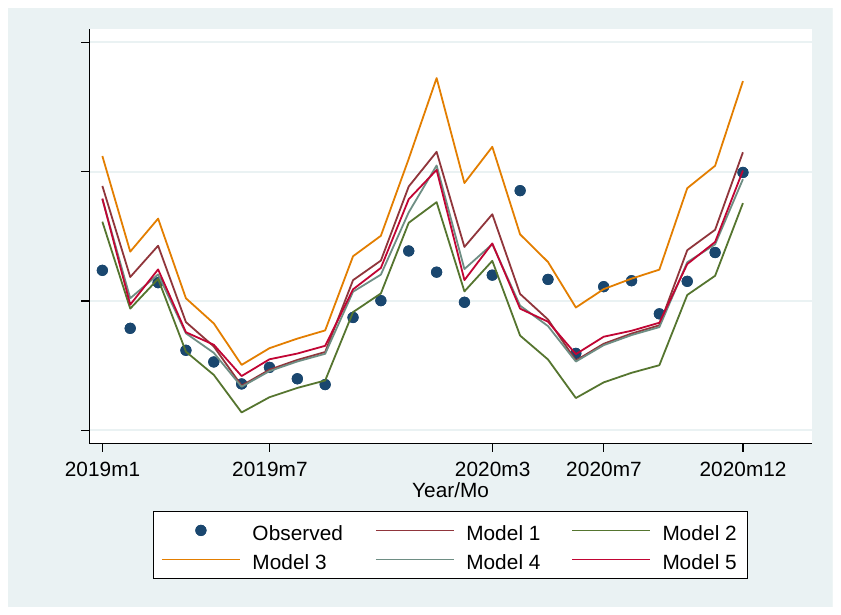

eFigure 12. Observed versus expected deaths from Alzheimer’s Disease during December among women aged 85 and older during 1999-2020

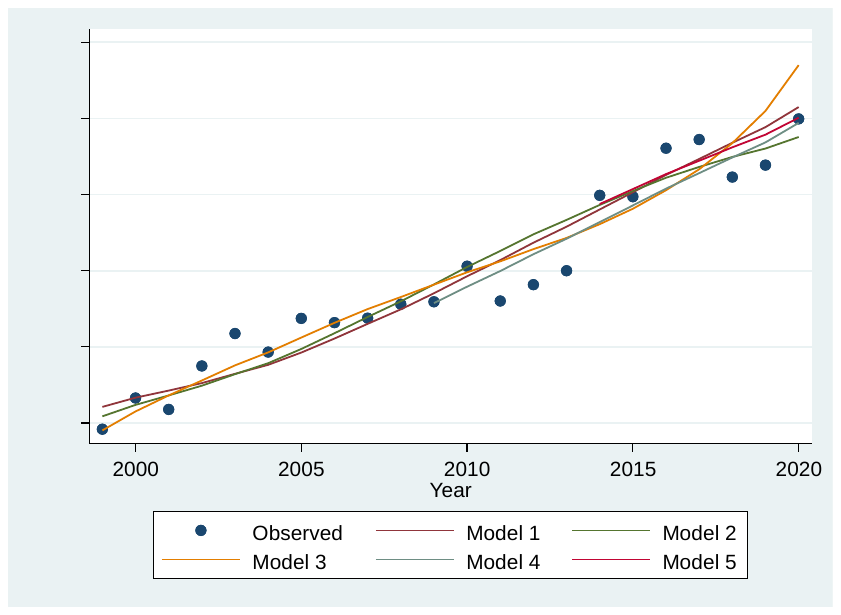

1. Currently, the provisional data by month, sex, and age group are available only for 13 broad groups of causes (including COVID-19). We grouped chronic lower respiratory diseases (J40-J47) and other respiratory diseases (J00-J06,J30-J39,J67,J70-J98) because the former accounted for vast majority of all deaths from respiratory diseases other than COVID-19 (78% in 2019 and 77% in 2020). There were relatively few deaths from the latter group, particularly below age 55.

   Initially, we also fit separate models for kidney disease (N00-N07, N17-N19, N25-N27), septicemia (A40-A41), and Ill-defined causes (R00-R99). The mortality rates from kidney disease, septicemia, and ill-defined causes were relatively low (even during the pandemic), particularly at younger ages. The results implied virtually no excess deaths from kidney disease (-518 for males, -413 for females) and septicemia (‑210 for males, 102 for females). Ill-defined causes contributed some excess deaths (i.e., 1,389 for males; 1,810 for females), but some of them could be COVID-19 deaths that were misclassified. A comparison of the number of deaths in that cause of death category across successive updates of the provisional data revealed that the category of ill-defined deaths may include some deaths that are later reclassified. The provisional data used for the main analysis^2^ was updated on July 27, 2021, whereas the less detailed dataset that comprised crude monthly death counts by selected causes of death was updated more than two months later on October 4, 2021.^3^ The death count for ill-defined causes in February 2021 declined 19% between the earlier and later files (late July vs. early October; 5 to 7 months after the death occurred), while the number of deaths from all-causes increased by 2% and the death count from external causes increased by 4%. Therefore, the results for these three groups of causes are not shown.
2. The provisional monthly death counts used in the main analysis^2^ include deaths from natural causes (ICD-10 A00-R99), but that data file does not include the death counts from external causes (V01-X59, Chapter XX of ICD-10), which represents a supplementary classification (<https://icd.who.int/browse10/2010/en#/XX>, accessed 9/10/2021). In most cases, deaths from Chapter XX were cross-classified in Chapter XIX (Injury, poisoning, and certain other consequences of external causes, S00-T98). For this analysis, the number of deaths from external causes is derived by subtracting deaths from natural causes from the death count for all causes.
3. As indicated by the 7-day COVID-19 death rates for the US as a whole,^6^ COVID-19 mortality rose rapidly between March 6, 2020 (0.01 per 100,000) and April 21 (4.87 per 100,000) and then generally declined until June 22 (1.29 per 100,000), thus ending the first wave. After that, the rates rose again, peaking on July 30 (2.55 per 100,000) and declining after that until September 30 (1.34 per 100,000), thus ending the second wave. Finally, the COVID-19 death rate increased again in late October through early January, peaking on January 13, 2021 (7.69 per 100,000) and fell after that to 0.41 per 100,000 on July 9, 2021. [A fourth wave peaked (at 3.87 per 100,000) on September 15, 2021.]
4. Cause of death certification for injury-related deaths often involve lengthy investigations (e.g., toxicology testing, autopsy).^10^
